# Deficiency of complement C3a and C5a receptors does not prevent angiotensin II–induced hypertension and hypertensive end-organ damage

**DOI:** 10.1101/2023.01.23.23284832

**Authors:** Marlies Bode, Georg Rudolf Herrnstadt, Leonie Dreher, Nicolas Ehnert, Pia Kirkerup, Maja T. Lindenmeyer, Catherine Meyer-Schwesinger, Heimo Ehmke, Jörg Köhl, Tobias B. Huber, Thorsten Wiech, Ulrich O. Wenzel

**Affiliations:** III. Department of Medicine, Cincinnati Children’s Hospital Medical Center, Cincinnati, Ohio, USA; Hamburg Center for Kidney Health (HCKH), University Medical Center Hamburg-Eppendorf, Cincinnati Children’s Hospital Medical Center, Cincinnati, Ohio, USA; Department of Cellular and Integrative Physiology University Hospital Hamburg-Eppendorf, Cincinnati Children’s Hospital Medical Center, Cincinnati, Ohio, USA; Institute for Systemic Inflammation Research, Lübeck, Germany, and Division of Immunobiology, Cincinnati Children’s Hospital Medical Center, Cincinnati, Ohio, USA; Department of Pathology, Section of Nephropathology, University Hospital Hamburg-Eppendorf, Hamburg, Germany

**Keywords:** C3aR, C5aR1, albuminuria, renal damage, cardiac damage, angiotensin II

## Abstract

Complement activation may drive the pathology of hypertension through its effects on innate and adaptive immune responses, aside from direct effects on tissue integrity. Recently it was suggested that hypertension is a disease characterized by a decreased number of forkhead box protein 3 (Foxp3)^+^ regulatory T cells (Tregs) and that combined deficiency of the anaphylatoxin receptors C3aR (complement component 3a receptor) and C5aR1 (complement component 5a receptor) upregulates Tregs and heals hypertension and hypertensive end-organ damage.

Using data from the European Renal cDNA Bank, renal single cell sequencing data and immunohistochemistry we found increased expression of C3aR, C5aR1 and Foxp3 in kidney biopsies of patients with hypertensive nephropathy. Expression of C3aR and C5aR1 was mainly found in myeloid cells and almost absent in lymphocytes. Next we aimed to determine whether C3aR or C3aR/C5aR1 double deficiency decreases blood pressure and hypertensive injury in Ang II infused mice.

However, no difference was found for blood pressure, renal injury (albuminuria, glomerular and tubulointerstitial injury, inflammation) and cardiac injury (cardiac fibrosis, heart weight, gene expression) between C3aR KO and WT mice as well as C3aR/C5aR1 double KO respectively. Ang II as well as DOCA salt induced hypertension resulted in an increased number of Tregs in the kidney. This was valid in mice of the Balb/c and C57black strain.

In summary, hypertensive nephropathy in patients is characterized by an increased expression of anaphylatoxin receptors and Tregs. However, deficiency of C3aR alone or C3aR and C5aR1 combined does not influence blood pressure or hypertensive end-organ damage in Ang II induced hypertension. Targeting the anaphylatoxin receptors C3aR alone or in combination with C5aR1 is not useful to treat cardiovascular disease in hypertension.

## Introduction

There is now overwhelming evidence that inflammation and immunity promote the development and maintenance of hypertension. (Drummond *et al*., 2019; Hengel *et al*., 2021; van der Heijden *et al*., 2022; Wenzel *et al*., 2016; Wenzel *et al*., 2020, Mathis *et al*., 2018). However, convincing therapeutic anti-inflammatory approaches to combat hypertension in patients are lacking. The immune system is divided into 2 branches, the innate and the adaptive. In this study we explore in humans and in mice the relationship between complement activation (an innate response) and regulatory T cells (Tregs) (an important player of adaptive response) to see whether targeting C3aR and C5aR1 could be a therapeutic option to treat hypertension in patients.

The complement system is an ancient part of innate immunity. It is central to the detection and destruction of invading microbes. It is constituted of a network of fluid-phase and cell membrane-bound proteins. Complement activation leads to generation of cleavage of highly active and proinflammatory fragments like the anaphylatoxins C3a and C5a. The role of anaphylatoxins in hypertension has been reviewed recently by us and others (Ruan *et al*., 2019; Wenzel *et al*., 2021c; Wenzel *et al*., 2017).

An important role for T cells in hypertension was shown in 2007, when it was found that the increase in blood pressure caused by Ang II infusion was significantly blunted in mice deficient for the recombinase-activating gene 1 (Rag-1^−/−^) which are lacking for T and B cells. However, the contribution of T cells to hypertension has recently been challenged by work from different groups including ours, that could not confirm increased resistance of Rag-1 deficient mice to Ang II infusion (Ji *et al*., 2017; Seniuk *et al*., 2020). An important subgroup of T cells are the regulatory T cells (Tregs), a subpopulation of CD4^+^ T cells with Foxp3 positivity. The main function of Tregs is maintenance of immunological tolerance. Several groups have shown that upregulation or adoptive transfer of Tregs lowers blood pressure and ameliorates cardiac and renal injury in different models of hypertension (summarized in Wenzel *et al*., 2016; Wenzel *et al*., 2021b).

The anaphylatoxins are known as regulators of Treg activity: C3a/C3aR and C5a/C5aR1 inhibit the function of CD4^+^ Foxp3^+^ circulating Tregs (Kwan *et al*., 2013), and absence of C3aR and C5aR1 signals in CD4^+^ T cells induces spontaneous Foxp3^+^ Treg differentiation upon T cells activation (Kwan *et al*., 2013).

Chen et al. have recently shown in an eloquent study that high blood pressure induced by Ang II infusion reduced the number of Foxp3^+^ Tregs in the kidney. Moreover, C3aR and C5aR1 double KO mice had an increased the number of Foxp3^+^ Tregs in the kidney and consequently revealed decreased blood pressure and hypertensive injury in response to Ang II, compared with wildtype mice (Chen *et al*., 2018). This is rather unusual since the number of Tregs in the kidney at least in a model of accelerated hypertension was increased in our own study (Krebs *et al*., 2014). Therefore, we got interested in the role of Tregs in hypertension and the possibility to upregulate them by double deficiency of C3aR and C5aR1 as a treatment option of hypertension.

To determine the role of Tregs in hypertension and the function of the C3aR/C5aR1 axis in hypertensive renal and cardiac injury we first examined the receptors and Tregs in kidney biopsies from hypertensive patients. Next, we first examined the effect of C3aR deficiency followed by C3aR/C5aR1 double deficiency in Ang II-induced hypertension.

## Methods

### Human Microarray Analysis

Human renal biopsy specimens were collected in an international multicentre study, the European Renal cDNA Bank-Kröner-Fresenius biopsy bank (ERCB-KFB, (Cohen *et al*., 2002). Biopsies were obtained from patients after informed consent and with approval of the local ethics committees. Following renal biopsy, the tissue was transferred to RNase inhibitor and microdissected into glomeruli and tubulointerstititum. Total RNA was isolated from microdissected glomeruli and tubuli, reverse transcribed, and linearly amplified according to a protocol previously reported (Cohen *et al*., 2006). Glomerular and tubular samples were analysed for mRNA expression levels. Analysis included gene expression profiles from patients with hypertensive nephropathy (glomeruli n=15, tubuli n=21), as well as controls (living donors, glomeruli n=41, tubuli n=42). CEL file normalization was performed with the Robust Multichip Average method using RMAExpress (Version 1.20) and the human Entrez-Gene custom CDF annotation from Brain Array version 25 (http://brainarray.mbni.med.umich.edu/Brainarray/Database/Custom/CDF_ download.asp). The log-transformed dataset was corrected for batch effect using ComBat from the GenePattern pipeline (http://www.broadinstitute.org/cancer/software/genepattern/). To identify differentially expressed genes, the SAM (Significance Analysis of Microarrays) method (Tusher *et al*., 2001) was applied using SAM function in Multiple Experiment Viewer (TiGR MeV, Version 4.9). A q-value below 5% was considered to be statistically significant.

### Kidney biopsies

From the archive of the institute of pathology, section for nephropathology of the University Hospital Hamburg-Eppendorf (UKE) biopsies of patients histopathologically diagnosed with so called malignant hypertension or stenosing arteriosclerosis were selected (n=13). The renal biopsy specimens were routinely examined by light microscopy, immunohistochemistry and electron microscopy. The diagnosis of malignant nephrosclerosis was defined as renal biopsy–proven vascular chronic changes: onion skin lesions, intimal fibrosis and arteriolar hyalinosis. Cases with clinical or serological evidence of malignancy, autoimmune disease, infection, drugs or pregnancy were excluded. Concurrent glomerulonephritis (GN) [e.g. immunoglobulin A (IgA) nephropathy] was also ruled out. For comparison, we took biopsies from patients with thin glomerular basement membrane disease (n=8). This is a non-immunologic mediated renal disease. So called zero-hour implantation biopsies of kidney allografts are not helpful since they may show complement activation due to the death of the donor. C3a-R (GTX114293, GeneTex) und C5AR1 (ABIN118843, antibodies-online.com) were used. For detection the ZytoChem Plus AP Polymer System POLAP-100 (Zytomed) was used. Approval by the ethics committee of the city of Hamburg (WF-011/15) was obtained.

### scRNA-seq Analysis

Preprocessed, quality controlled scRNA-seq data of 51,849 CD10^negative^ single cells from kidney biopsies of patients with hypertensive nephropathy or healthy controls were obtained from Kuppe et al. published in Nature recently (Kuppe *et al*., 2021) and re-analyzed using scRNA-seq analysis R package Seurat (version 4.1.0). More than 80% of renal cortical cells are proximal tubular epithelial cells and are dominating single cell maps and masking other populations. Therefore, analysis was performed in a data set using viable, non-proximal tubular cells sorted for (CD10^negative^). CD10 was used as a marker of proximal tubule cells. However, also CD10^negative^ cells contain proximale tubule cells. To analyze gene expression of regulatory T cells in healthy kidneys with re-analyzed our own data of 5905 single CD3^+^ T cells obtained from three tumor nephrectomies (Krebs et al., 2020). We used LogNormalize method in Seurat to normalize the scRNA-seq data before scaling by ScaleData function (default parameters). Highly variable genes were identified by function FindVariableFeatures, selection.method = "vst", nfeatures = 2000. Principal component analysis was performed on the scaled data (function RunPCA, npcs = 50) to reduce dimensionality. Principal components were used to compute the KNN graph based on the euclidean distance (function FindNeighbors) to subsequently generate cell clusters using function FindClusters. Uniform Manifold Approximation and Projection (UMAP) or t-Distributed Stochastic Neighbor Embedding (tSNE) was used to visualize clustering results. The top DEG in each cluster were found using the FindAllMarkers function (min.pct = 0.25, logfc.threshold = 0.25) that ran Wilcoxon rank sum tests. The top marker genes were then compared and validated with the original publication and used to determine the cell types of each cluster. Seurat functions AverageExpression and DoHeatmap were used to visualize cluster identifying marker genes for each cell cluster. Gene expressions were visualized using Seurat functions FeaturePlot and DotPlot. Disease condition stratified gene expressions were visualized using Seurat function VlnPlot and modified by python package Seaborn (version 0.11.2).

### Mice and experimental groups

C3aR^-/-^ and C5aR1^-/-^ mice were obtained from J. Koehl, Lübeck Germany. C3aR/C5aR1 double deficient mice were constructed by breeding C3aR^-/-^ with C5aR1^-/-^ mice. Matched WT mice on the Balb/c background were used. WT mice were randomly distributed between control and hypertensive group, while every KO mouse was allocated to hypertension. All animal experiments were approved by local ethic committee and local animal authority. Handling of mice was not done in a completely blinded fashion since normotensive and hypertensive animals are easily distinguished. However, hypertensive WT and KO mice were not specifically labelled. For detection of C3aR on leukocytes and resident renal a floxed tandem-dye Tomato (tdTomato)–C3aR reporter knock-in mouse was used (Quell *et al*., 2017). For isolation of Tregs BAC-transgenic Rorc(gt)-Gfp^TG^ x Foxp3-IRES-mRFP (FIR) mice were kindly provided by Gérard Eberl, Paris, France and Matthias Lochner, Hannover, Germany.

### Ang II-induced hypertension

Due to the fact that mice of the C57black strain are resistant to hypertensive renal injury by Ang II infusion (Wesseling *et al*., 2005), we induced an accelerated model of hypertension as previously described by us (Ahadzadeh *et al*., 2018; Weiss *et al*., 2016). C3aR^-/-^ and C5aR1/C3aR^-/-^ double KO mice have no renal phenotype at baseline. Therefore, normotensive C3aR^-/-^ and C5aR1/C3aR^-/-^ mice were not examined in the current experiment to reduce the number of mice as defined by ARRIVE guidelines. For the induction of hypertension mice, aged 10 weeks were uninephrectomized on day 0. Anaesthesia was given by 4% isoflurane and analgesia by 0.1 mg/kg buprenorphine s.c.. Mice received 0.9% NaCl in the drinking water starting on day 7 followed by constant administration of 0.75 ng of Ang II (Bachem, Switzerland) per gram body weight per minute through subcutaneously implantation of an osmotic minipump (Alzet 1002, Cupertino, USA) on day 14. Mice were sacrificed at day 28, 2 weeks after commencement of Ang II infusion, as described by us and depicted in supplement figure 1 (Ahadzadeh *et al*., 2018; Weiss *et al*., 2016).

### DOCA salt induced hypertension

At day 0 unilateral nephrectomy was performed in mice aged 10 weeks. A deoxycorticosterone acetate (DOCA) pellet (50 mg, 21-day release, Innovative Research of America, USA) was implanted subcutaneously, and 0.9% NaCl added to the drinking water. Anaesthesia was given by 4% isoflurane and analgesia by 0.1 mg/kg buprenorphine s.c.. After 21 days, the DOCA pellet was renewed, after 42 days the mice were sacrificed, and organs removed as depicted in supplement figure 1.

### Systolic blood pressure

Systolic blood pressure was measured with a computerized tail cuff system (MC4000, Hatteras Instruments, USA,) as described (Ahadzadeh *et al*., 2018; Weiss *et al*., 2016). With appropriate training and device tail cuff measured blood pressure is reliable (Kurtz *et al*., 2005).

### Glomerular filtration rate (GFR) measurement

GFR was measured transcutaneously in conscious mice. The technique relies on a miniaturized device equipped with an internal memory that permits the transcutaneous measurement of the elimination kinetics of the fluorescent renal marker FITC-sinistrin. The device is started to measure the background signal for 1 min before the FITC-sinistrin is given by retrobulbar injection. The measurement runs for one hour while mice are awake and have free access to food and water. GFR is calculated from FITC-sinistrin plasma clearance with „MPD lab“ software (Schreiber *et al*., 2012).

### Plasma and urine analysis

Mice were placed into metabolic cages for a 6-hour urine collection. At the end of the experimental period, heparinized blood was collected. Plasma cholesterol and urine creatinine were measured by an autoanalyzer (Ahadzadeh *et al*., 2018; Weiss *et al*., 2016). Albumin was measured by ELISA (Bethyl Laboratories, Montgomery, USA). Albumin/creatinine ratio was calculated to assess the extent of albuminuria.

### Histopathologic analysis

After removal of kidney and heart, tissues were fixed with 4% neutral buffered formalin, embedded in paraffin and sectioned at 1 μm thickness. Sections were then deparaffinized and stained for light microscopy with PAS (renal tissue) or Sirius Red Fast Green (SRFG, cardiac tissue). Glomerular injury was evaluated histologically using a semi-quantitative scale with 0 indicating normal architecture, 1 mild injury in less than a third of the glomerular tuft (small mesangial expansion with matrix, cells and sclerosis), 2 damage of more than a third of the glomerular tuft, and 3 damage of the whole glomerulus. Under 400-x magnification, 30 glomeruli per animal were analysed (Krebs *et al*., 2014). The glomerular size was measured by computer-aided assessment. Tubulointerstitial damage was assessed by using a semi quantitative scale with 0 indicating no injury, 1 mild injury, 2 moderate, and 3 intensive injury. Injury was defined by widening of tubular lumen, flattening and atrophy of tubular cells with enlargement of the basement membrane and widening and scarring of the interstitial space. Under 400x magnification 30 fields per kidney were analyzed (Ahadzadeh *et al*., 2018; Weiss *et al*., 2016). Cardiac fibrosis was evaluated by scoring heart sections using a scoring from 0 to 3 as described by us (Krebs *et al*., 2012). Ratings covered values between categories. Scoring was done in a blinded fashion.

### Real-time quantitative RT-PCR

Total RNA from kidney cortex or heart ventricles was isolated using the RNeasy kit (Qiagen,Netherlands). Real-time quantitative PCR was performed using Thermo Fisher Scientific’s Quantstudio 3 system and PowerUp SYBR Green Mastermix (Thermo Fisher Scientific, USA). Mouse-specific PCR primers were used. The levels of mRNA expression in each sample were normalized to 18S rRNA expression (Krebs *et al*., 2012; Lehners *et al*., 2014).

### Flow cytometry

Kidneys were minced and incubated in digestion medium (RPMI 1640 containing 10% FCS, 1% penicillin/streptomycin, 1% HEPES, 0.1mg/ml DNAse and 0.4mg/ml Collagenase D).To obtain a single-cell solution, kidneys were dissociated using the gentleMACS Dissociator. A Percoll gradient was used to further purify viable cells from cell debris. Erythrocytes were lysed with ammonium chloride. For flow cytometry, cells were surface-stained with fluorochrome-conjugated antibodies: CD45 (30-F11), CD3 (17A2), γδTCR (Gl3), CD4 (RM4- 5), CD8 (53-6.7), CD11b (M1/70), Ly6G (1A8), CD11c (N418/HL3), MHCII (M5/114.15.2), Ly6C (AL-21/HK1.4) and F4/80 (BM8) (Biolegend, San Diego, USA, and BD Biosciences, Franklin Lakes, USA). Dead cells were excluded during flow cytometry via LIVE/DEAD staining (Invitrogen Molecular Probes, Carlsbad, USA). After permeabilisation the cells were stained for the transcriptional factors RORγt (Q31-378), Foxp3 (FJK-16s), Tbet (4B10) and GATA3 (L50-823) (BD Biosciences) and fixated (Foxp3/Transcription Staining Buffer Set) (Invitrogen). Data analysis was performed with FlowJo (Tree Star, USA).

### Statistics

For the statistical analysis, GraphPad Prism 8.3.0 was used. The Kolmogorov–Smirnov test was used to test for normal distribution of the values. In case of normal distribution, for multiple comparisons one-way analysis of variance (ANOVA) and post hoc analysis by Tukey’s multiple comparisons test with a single pooled variance were performed. If data were not normally distributed, Kruskal–Wallis with Dunn’s multiple comparisons test was used. For comparison of two parameters, unpaired t-test for normally distributed and Mann–Whitney test for not normally distributed parameters were used. Survival percentages were calculated by Kaplan-Meier and survival curves compared with Log-rank (Mantel-Cox) test and Log-rank test for trend. Differences were considered statistically significant at * p<0.05, **p<0.01, ***p<0.001, and ****p<0.0001

## Results

### C3aR, C5aR1 and Foxp3 in patients with hypertensive nephropathy

For analysis of anaphylatoxin receptors and Foxp3 in hypertensive patients we took advantage of the European renal cDNA bank (ERCB). We examined expression in renal biopsies from patients with hypertensive nephropathies (n=21) and compared these data with kidney biopsies from living donors (n=42). Expression data are available for microdissected glomeruli and tubulointerstitium. C3aR expression was significantly increased in glomeruli from hypertensive patients and no effect was seen in the tubulointerstitial space. No glomerular induction was found for C5aR1 and its tubulointerstitial expression level was below detection limit. A small but significant increase was found for Foxp3 in the tubulointerstitium (figure 1A).

**Figure 1.**
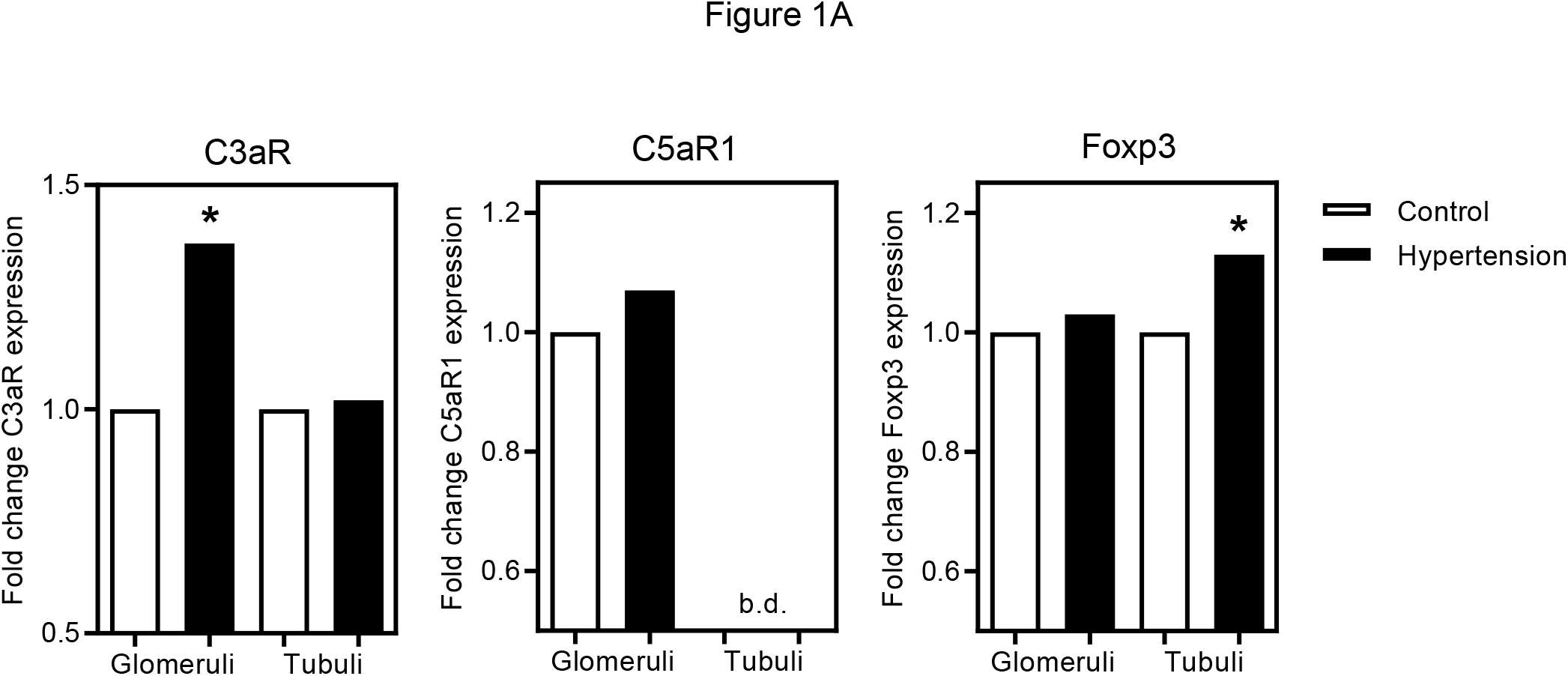
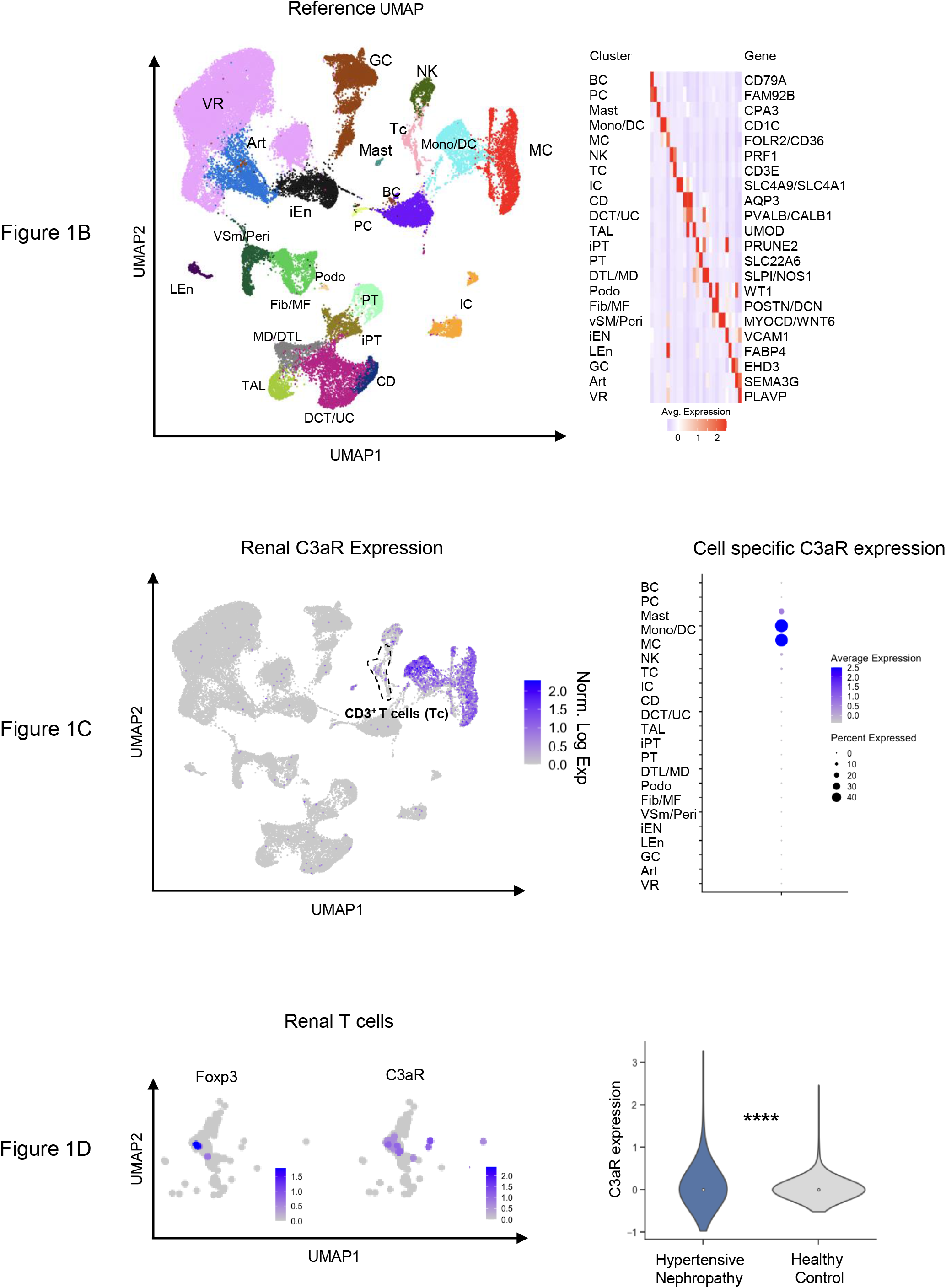
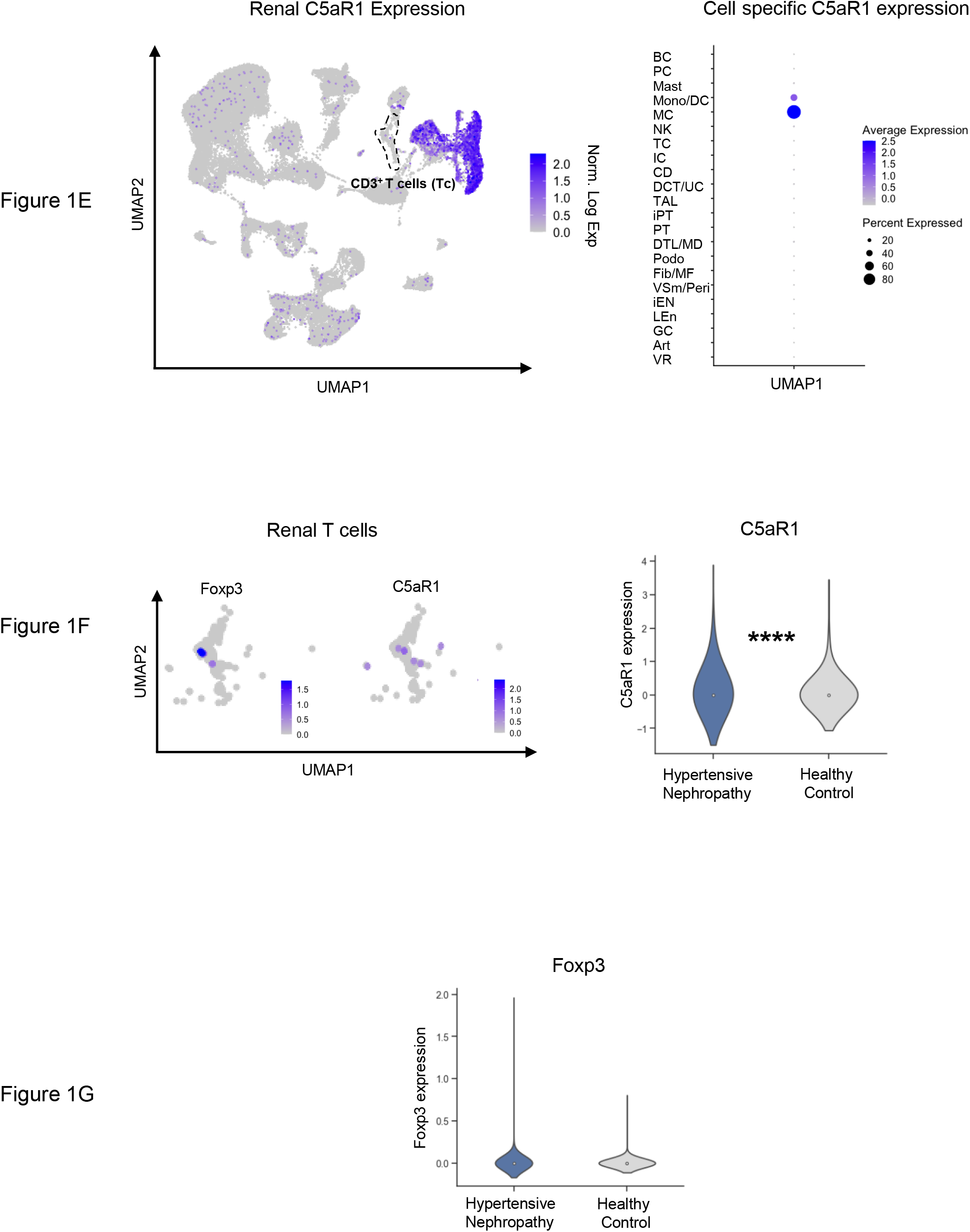
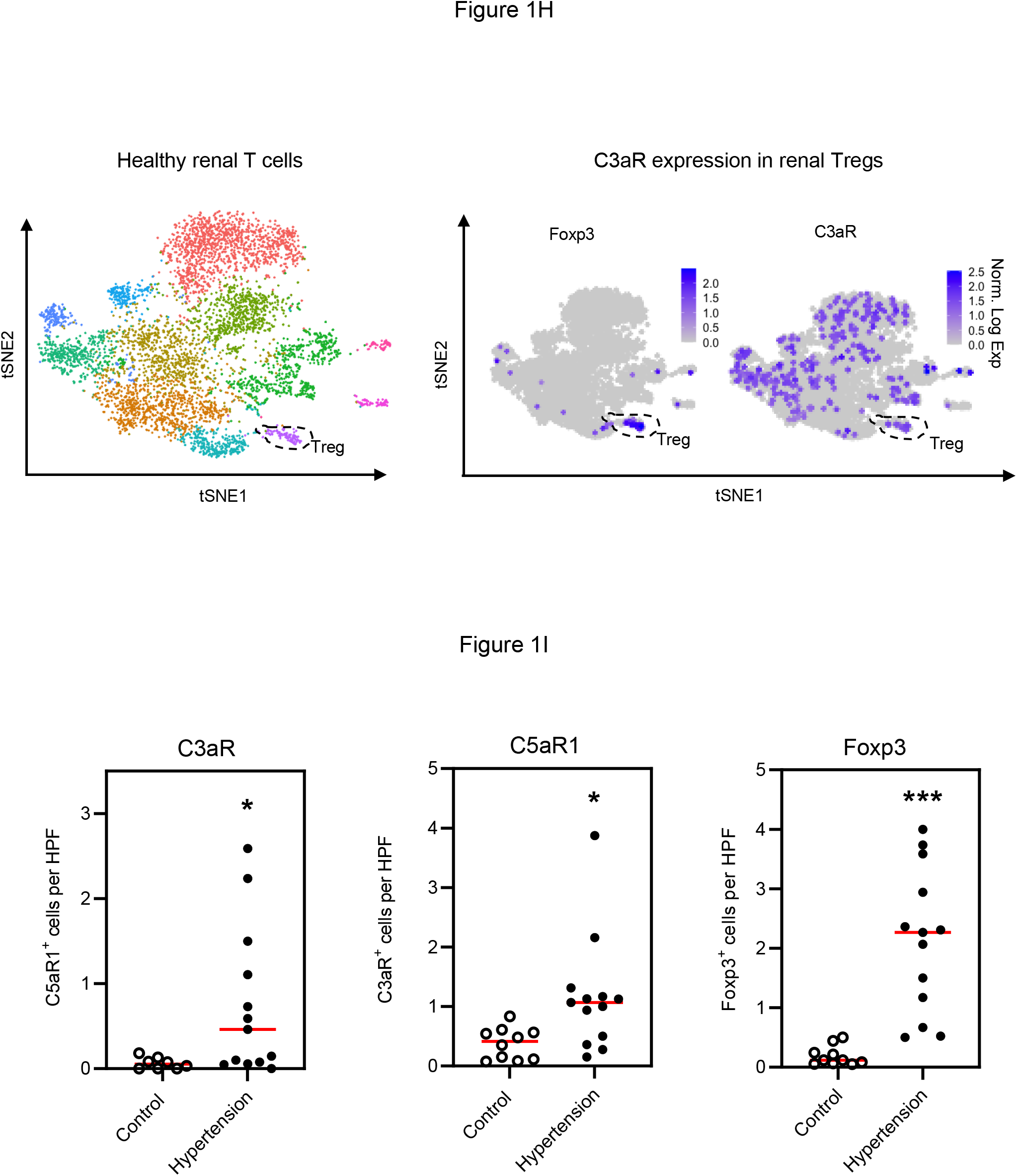
Tregs in human kidney biopsies. A: Gene expression analysis revealed significantly increased glomerular C3aR and tubulointerstitial Foxp3 expression in renal biopsies from patients with hypertensive nephropathies compared to kidney biopsies from living donors. B: Left panel: Re-analyzed reference UMAP embedding of 51,849 CD10^negative^ single cells from 15 human kidneys. BC, B cells; PC, plasma cells; Mast, mast cells; Mono/DC, monocytes/dendritic cells; MC, macrophages; NK, NK cells; TC, T cells; IC, intercalated cells; CD, collecting Duct; DCT/UC, distal convoluting tubule/urothel cells, TAL, thick ascending limb; iPT, injured proximal tubule, PT, proximal tubule; DTL/DM, distal convoluted tubule/macula densa; Podo, podocytes; Fib/MF, fibroblast/myofibroblast; vSM/Peri, vascular smooth muscle cell/pericytes; iEN, injured endothelium; LEn, lymph endothelium, GC, glomerular capillaries; Art, arteriolar endothelium; VR, vasa recta. Right panel: Scaled gene expression of top cluster identifying genes. Each column is the average expression of all cells in a cluster. C: UMAP plot showing C3aR gene expression (scale bars indicate normalized expression). T cell cluster surrounded by dashed line. Right panel: Dot plot of C3aR gene expression in each cluster. The size of the dot denotes the percentage of cells in each cluster expressing C3aR; the intensity of color represents mean gene expression. D: UMAP plot of scaled up T cell cluster showing Foxp3 and C3aR expression. Right panel: Violin plot of C3aR expression in 51,849 CD10^negative^ single cells stratified by disease condition. E: UMAP plot showing C5aR1 gene expression (scale bars indicate normalized expression). T cell cluster surrounded by dashed line. Right panel: Dot plot of C5aR1 gene expression in each cluster. The size of the dot denotes the percentage of cells in each cluster expressing C5aR1; the intensity of color represents mean gene expression. F: UMAP plot of scaled up T cell cluster showing Foxp3 and C5aR1 expression. Right panel: Violin plot of C5aR1 expression in 51,849 CD10^negative^ single cells stratified by disease condition. G: Violin plot Foxp3 expression in 51,849 CD10^negative^ single cells stratified by disease condition. H: tSNE (t-distributed stochastic neighbor embedding**) p**lot showing unsupervised clustering of 5905 single CD3^+^ T cells of healthy areas from tumor nephrectomies (n=3). Purple cluster represents regulatory T cells, surrounded by dashed line. tSNE plot showing Treg identifying Foxp3 and C3aR gene expression (scale bars indicate normalized expression). Treg cluster surrounded by dashed line. I: Number of C3aR, C5aR1 and Foxp3^+^ cells/high power field in renal cortex of controls and patients with hypertension. *=p<0.05 ***=p<0.001 ****=p<0.0001 controls vs. hypertensive nephropathy.

After analysing the glomerular and the tubulointerstitial compartment we next examined C3aR and C5aR1 expression in a single cell RNAseq approach by re-analyzing an integrated scRNA-seq dataset of renal biopsies from healthy controls and patients with hypertensive nephropathy (Kuppe *et al*., 2021). Figure 1B shows the different cell compartments and their defining genes. Analysis revealed C3aR and C5aR1 expression in different cell types in the kidney (figure 1C and E). C3aR and C5aR1 expression was mainly detected in monocytes, dendritic cells and macrophages. T cells in contrast showed only minor, but detectable expression of both genes (figure 1D and F). C3aR and C5aR1 expression was significantly higher in hypertensive nephropathy compared to healthy controls, although mean differences between the groups were low (figure 1D and F). Gene expression of regulatory T cell main transcription factor FOXP3 was found in parts of the T cell cluster, showing overlap expression with C3aR but not for C5aR1 (figure 1D and F). No difference was found between Foxp3 expression between normotensive and hypertensive patients (figure 1G).

To verify whether renal Tregs express C3aR or C5aR1 we next analyzed our own data set of T cells isolated from healthy kidneys. Unsupervised clustering revealed a small Treg cluster confirming robust but low C3aR expression (figure 1H). C5aR1 expression however could not be detected (data not shown).

Quantification of C3aR, C5aR1 and Foxp3 positive cells by immunohistochemistry in controls and patients with hypertensive nephropathy showed a significant increased number of positive cells in the renal cortex in hypertension (figure 1I).

### C3aR in mice

First we determined which cells in the kidney express the C3aR. We took advantage of the recently developed tdTomato-C3aR knock-in mouse, in which all C3aR-expressing cells are tomato-positive (Karsten *et al*., 2015). Using confocal microscopy for tomato expression in renal tissue we found that almost exclusively tomato-positive cells in the kidney were found in a periglomerular and interstitial localization. Shape, form and localization of these cells suggested that these cells were dendritic cells and/or monocytes/macrophages (figure 2A). Resident renal cells showed no tomato-positivity. Next we asked which infiltrating cells in the kidney are expressing the C3aR. Leukocytes were isolated from the kidney and examined by FACS analysis. The two gating strategies are shown in suppl. figure 2A and B. FACS analysis revealed that 27.4% of CD45 positive cells expressed C3aR as shown in figure 2C. The majority of these positive cells were dendritic cells (96.0%) followed by macrophages (3.4%) and neutrophils (0.3%) as shown in figure 2C and figure 2D. Additionally, we analyzed the frequency of C3aR-positive cells within the different leukocytes (Figure 2E) and especially myeloid cell populations as shown in figure 2F. We found that 95.0% of dendritic cells in the kidney were C3aR positive. 12.7% of macrophages and 1.2% of neutrophils expressed the receptor (FACS blots figure 2F, quantification in 2G). We also examined lymphocytes. Gating for CD3 and tomato revealed that only very few lymphocytes were positive for C3aR. 0.6% of lymphocytes were tomato positive as shown in suppl Figure 2C and D. Very few CD4^+^ cells were positive and no staining was found in γδ T cells and CD8^+^ T cells. Additionally, we analyzed the frequency of C3aR-positive cells within the different lymphoid cell populations (suppl. figure 2E and F). 0.6% of CD3^+^ cells and 0.2% of CD4^+^ T cells expressed C3aR. No C3aR staining was found in γδ and CD8^+^ T cells. Taken together the human and murine data show that C3aR in the kidney is mainly expressed in dendritic cells. Monocytes/macrophages and neutrophils express to a certain percentage the receptor, and lymphocytes express only a negatable amount of it. No expression is found on resident renal cells.

**Figure 2.**
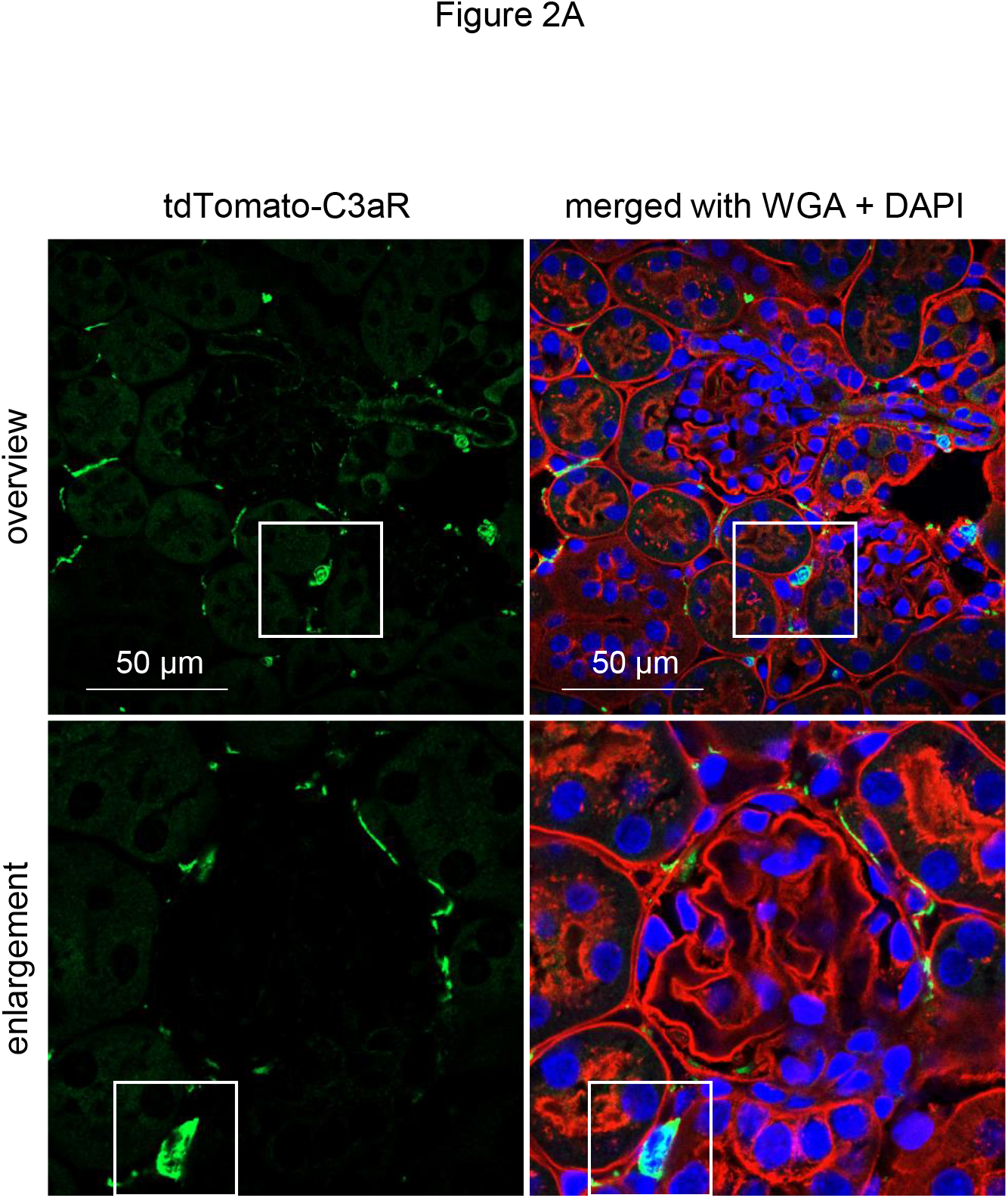
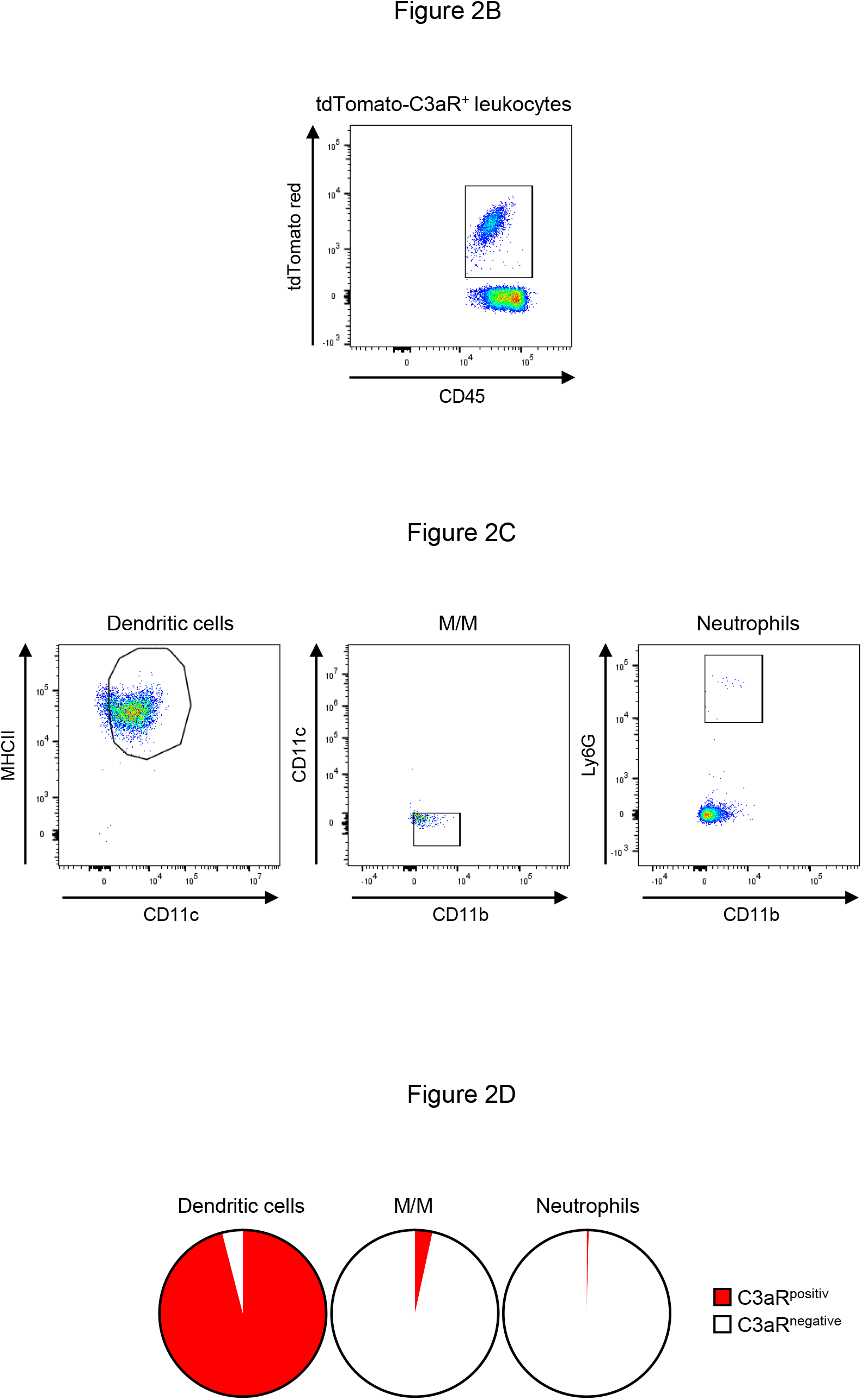
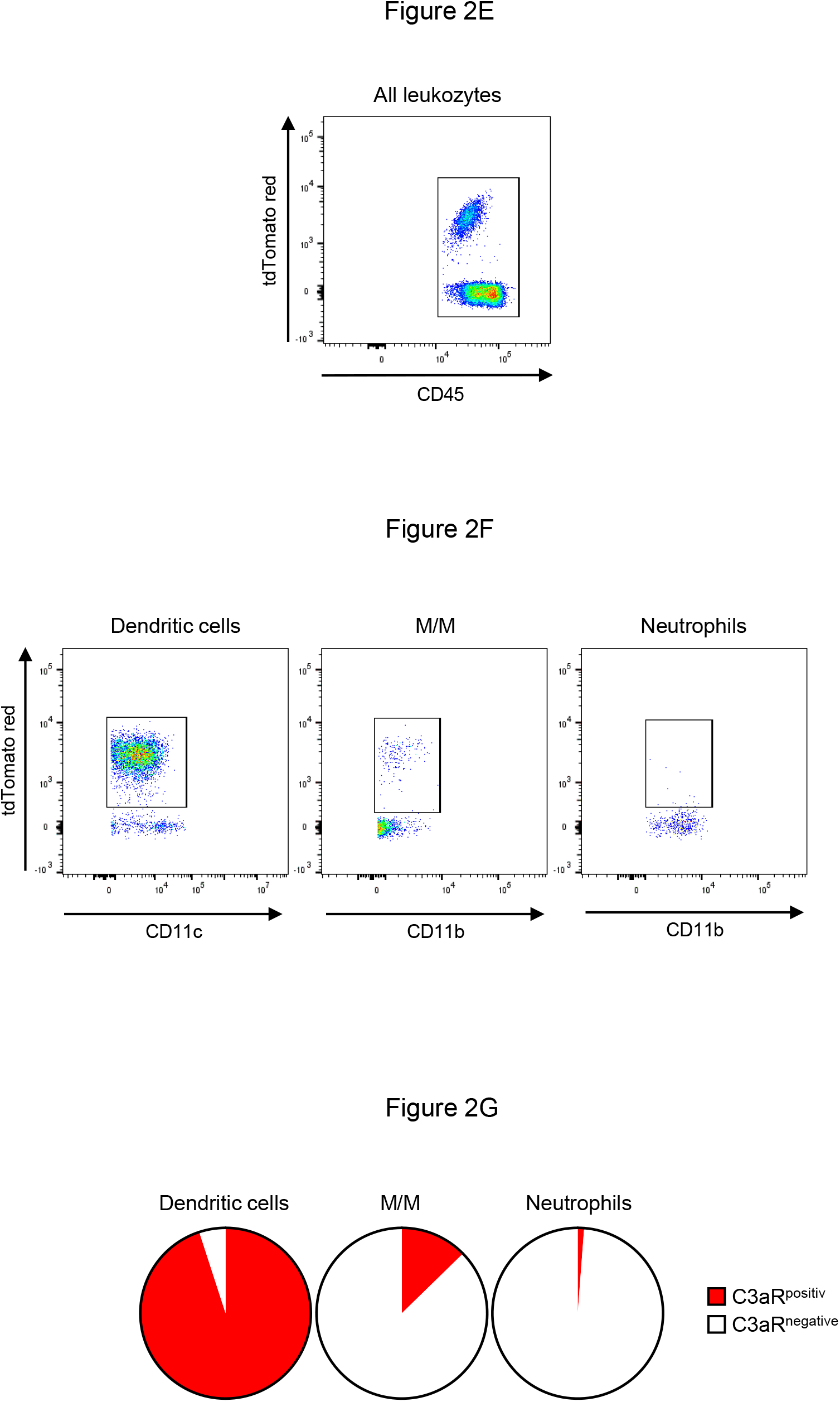
C3aR. A: Confocal microscopy for tomato expression in renal tissue showed tomato-positive cells in in a periglomerular and interstitial localization. Shape, form and localization of the cells suggested that these cells were dendritic cells and/or monocytes/macrophages. WGA wheat germ agglutinin DAPI 4′,6-diamidino-2-phenylindole. B: FACS analysis revealed that up to 27.4% of the infiltrating CD45 positive cells were C3aR positive. C and D: This population contains of 96.0% dendritic cells, 3.4% macrophages and 0.4% neutrophils. E: Gating CD45 positive cells and all leukocyte populations first and afterwards the C3aR reporter positive cells. F and G: 95% of the dendritic cells, 12.7% of the macrophages and 1.2% of the neutrophils are positive for C3aR.

### Hypertension and renal injury C3aR KO

We next examined the effect of C3aR deficiency on Ang II induced hypertension and end-organ damage. Ang II infusion increased systolic blood pressure in WT and C3aR-deficient mice with no difference between the two groups as shown in figure 3A. We also calculated the area under the curve for systolic blood pressure and found no difference between WT and KO mice (figure 3A). Mortality was found in hypertensive mice without a significant difference between both genotypes (16 of 34 in WT, 25 of 45 KO).

**Figure 3.**
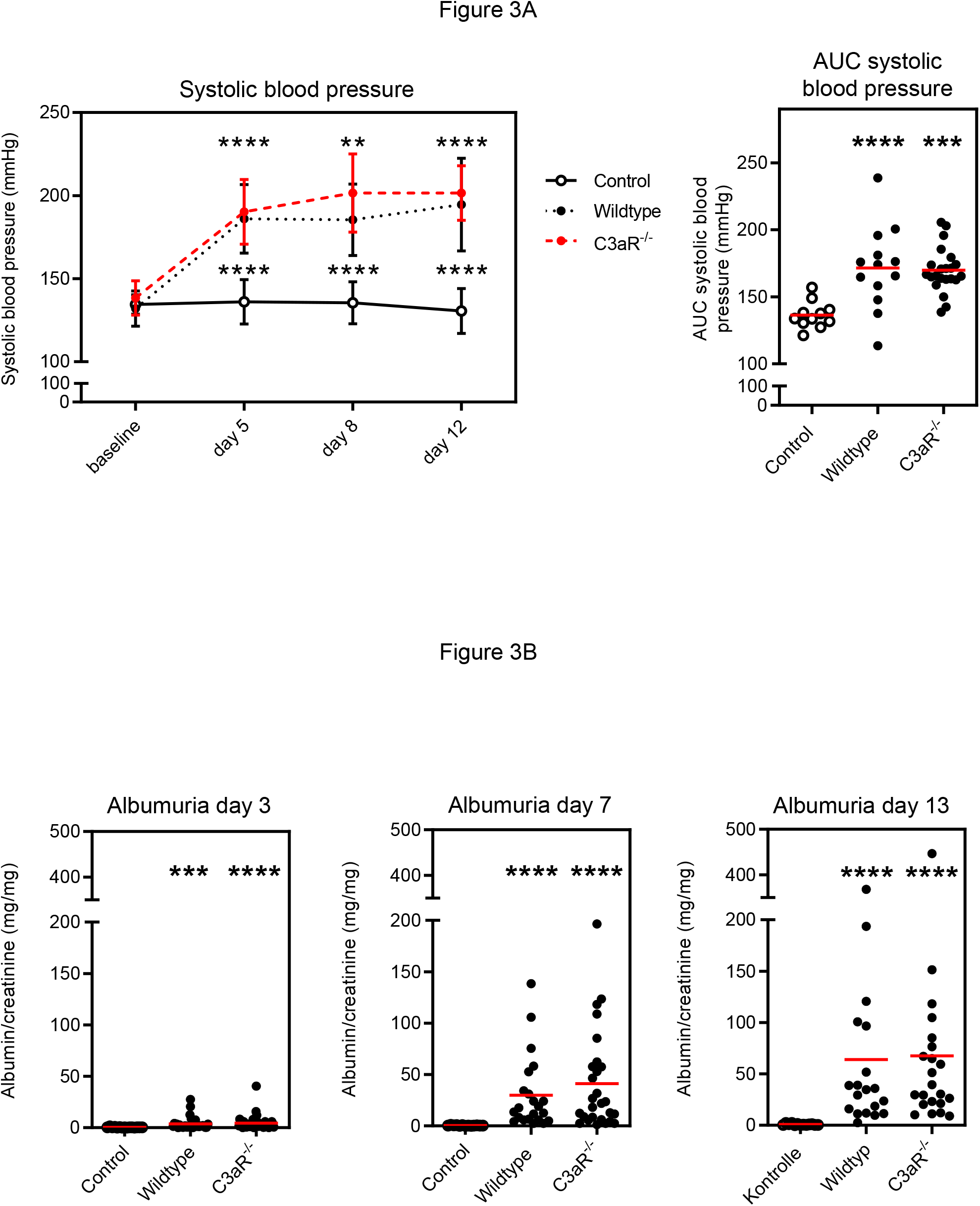
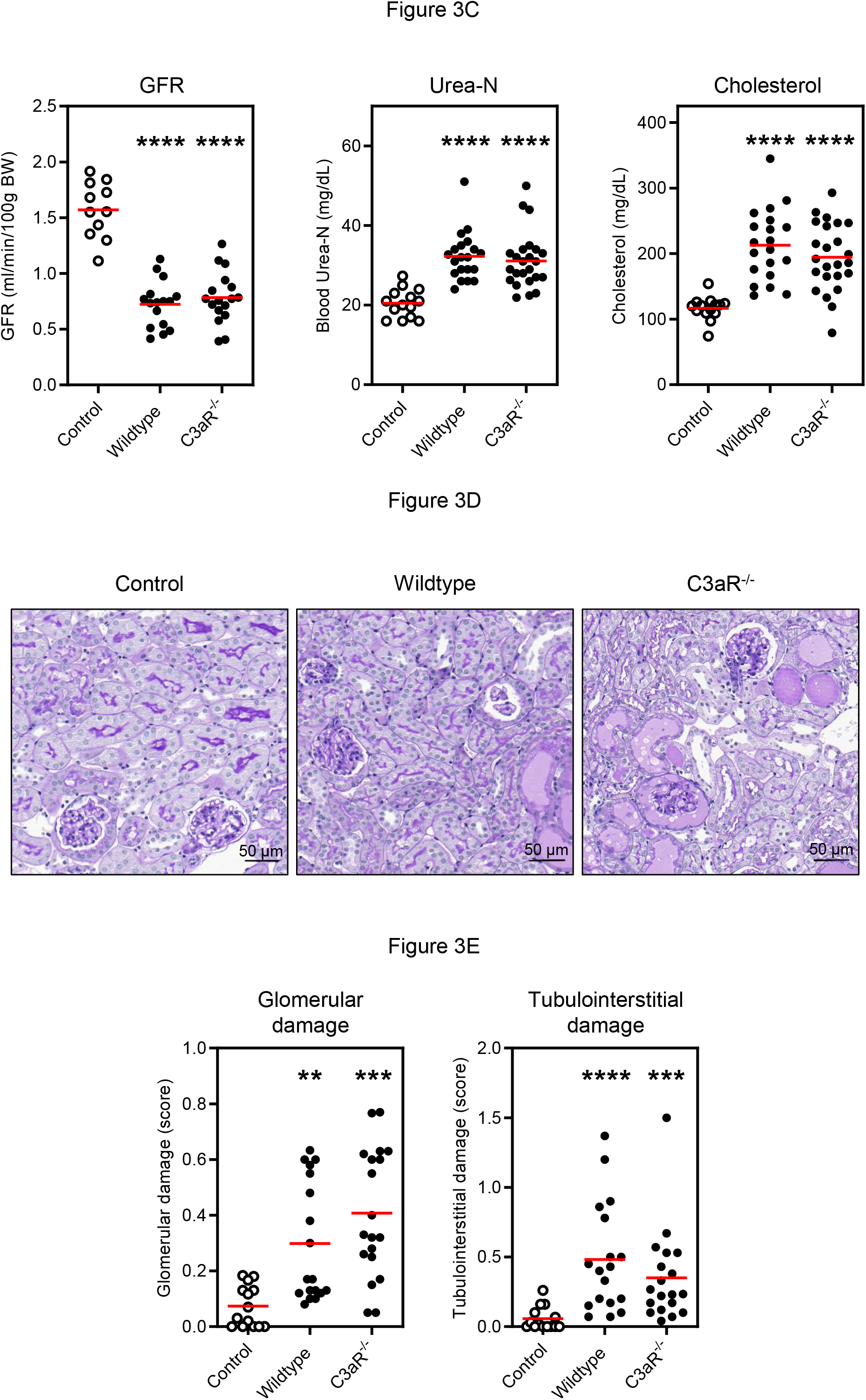
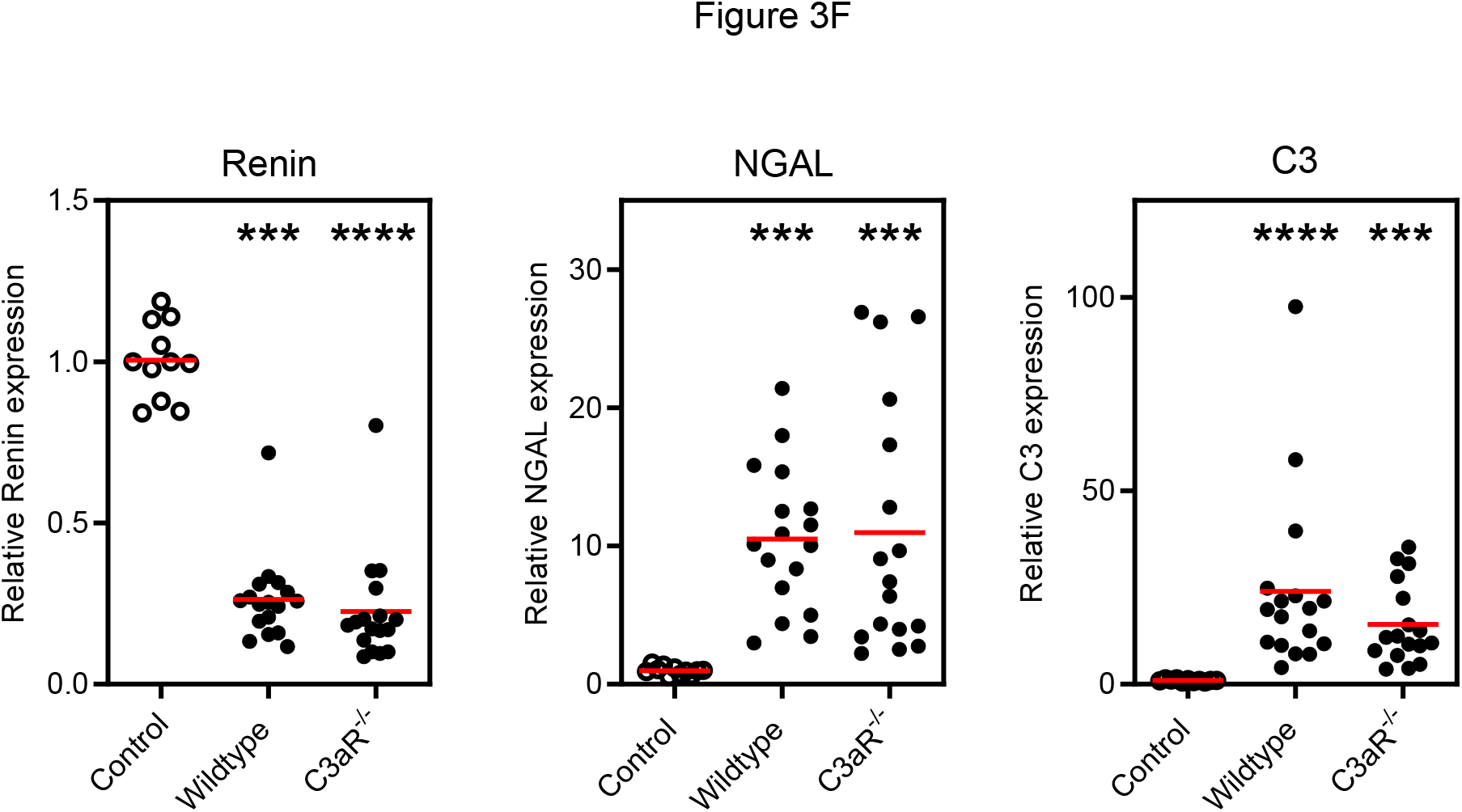
Systolic blood pressure and renal injury C3aR. A: Ang II infusion increased systolic blood pressure in WT and C3aR KO mice during the different time points of the experiment with no difference between the two groups. This was also valid for the area under the curve (AUC) of blood pressure. B: Ang II induced heavy albuminuria without difference in both genotypes. C: Ang II decreased GFR in both groups. Accordingly, plasma values of urea-N were significantly elevated. Plasma values of cholesterol were also significantly increased in both hypertensive groups. D: PAS stained sections revealed glomerular changes with mesangial expansion and sclerosis in both genotypes. E: Scoring of glomerular and tubulointerstitial damage showed no significant differences between WT and C3aR-deficient mice. F: Suppression of renal renin and increased expression of NGAL as well as C3 were found in both hypertensive groups. White dots normotensive mice, black dots hypertensive mice. **=p<0.01 ***=p<0.001, ****=p<0.0001 vs. normotensive control

No albuminuria was found in C3aR deficient mice at the beginning of the experiment indicating that these mice do not have a basal renal phenotype or renal abnormalities. Ang II induced heavy albuminuria in WT and C3aR-deficient mice on day 3, 7 and 13 without difference between both genotypes as shown in figure 3B. For a detailed analysis of kidney function, we measured the GFR in a subset of mice by using a FITC-sinistrin based technique, which allows measurement in conscious mice. First, we performed a basal measurement before induction of hypertension. GFR averaged 1.53±0.05 in WT and 1.52±0.07 ml/min/100 g body weight in KO mice showing no difference in renal function before the start of the experiment. This confirms that C3aR^-/-^ mice do not have a renal phenotype. Ang II administration caused a drop of GFR in hypertensive mice at the end of the experiment (46% of normotensive controls in WT and 50% in C3aR-deficient mice) without difference between both genotypes (figure 3C). Plasma urea is another marker of renal function and was significantly elevated in Ang II infused mice on day 14 without difference between WT and KO mice (Figure 3C). Heavy albuminuria induces a nephrotic syndrome with elevated cholesterol levels. Plasma levels of cholesterol were increased in both hypertensive groups. Next, we examined the kidney histology. Normal renal architecture was detected in normotensive controls. Ang II infusion induced hypertensive glomerular injury with mesangial expansion and sclerosis in WT and C3aR^-/-^ mice shown in figure 3D. Semi-quantitative analysis of glomerular injury by scoring revealed similar glomerular changes in C3aR-deficient and WT mice. Similarly, no difference between both genotypes was found for tubulointerstitial damage (figure 3E). Additionally, we utilised real-time PCR analysis to quantify genes of renal injury. Real time PCR analysis showed a similar decreased expression of renin in both groups in response to Ang II infusion. In addition, NGAL, a marker or renal injury, was significantly upregulated but no difference was found between both hypertensive groups. C3 is the central protein of the complement cascade and C3a is derived from it. Interestingly, Ang II infusion increased significantly renal expression of C3 in both genotypes (figure 3F).

Next we analysed leukocytes infiltrating the kidney in hypertension in the presence or absence of C3aR in a subset of mice. Gating strategy is shown in suppl. figure 3A. Since dendritic cells are the main C3aR expressing cell population in the kidney we first evaluated these cells. There was a slight but not significant increase in hypertensive WT and KO mice and no difference between both genotypes (suppl. figure 3B). Also, the frequencies of neutrophils (PMNs) and macrophages within the CD45^+^ kidney cells were modestly increased and similar in hypertensive WT and C3aR-deficient mice (suppl. figure 3B). In addition, the frequencies of CD3, CD4 and CD8 were not different between both genotypes in hypertension. C3aR deficient mice had a significant increased number of γδ T cells infiltrating the kidney. The frequencies of Th1, Th17 and Tregs cells were indistinguishable between hypertensive WT and C3aR-deficient mice (suppl. figure 3C).

### Cardiac injury C3aR KO

Next we examined cardia injury. No fibrosis was detected in normotensive control mice as shown in figure 4A. Ang II infusion induced extensive cardiac fibrosis. Scoring of the fibrosis revealed no difference between both genotypes (figure 4B). Cardiac hypertrophy is an important predictor of hypertensive end-organ damage. Relative heart weight was not different between C3aR-deficient and WT mice (figure 4B). The presence of a significantly increased gene expression of ANP and BNP in both genotypes further supports a similar cardiac injury (figure 4C). Taken together these results endorse similar cardiac injury in WT and C3aR KO mice.

**Figure 4.**
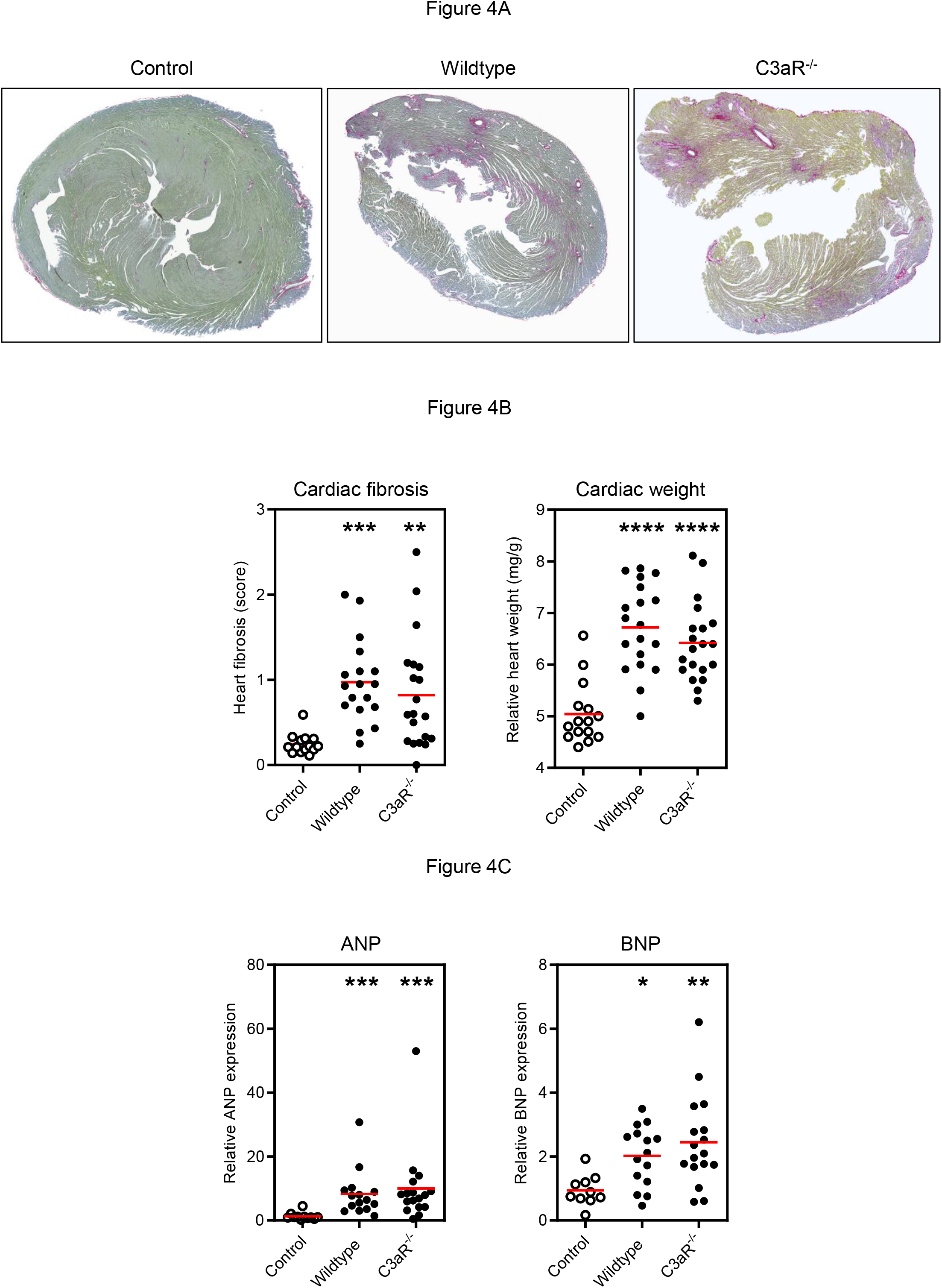
Cardiac injury C3aR. A: No fibrosis was seen in normotensive controls. Ang II-induced extensive cardiac fibrosis as shown in Sirius Red Fast Green stained cardiac sections. B: Cardiac fibrosis measured by scoring was not different between WT and KO mice. Relative heart weight was increased in hypertension with no difference between both groups. C: Hypertension induced increased cardiac expression of ANP, BNP and no difference was found between both genotypes. White dots normotensive mice, black dots hypertensive mice. *=p<0.05, **=p<0.01, ***=p<0.001, ****=p<0.0001 vs. control

### Hypertension and renal injury C3aRxC5aR1 double KO

We next examined whether double KO of C3aR and C5aR1 ameliorates hypertension and end-organ damage. Ang II infusion increased systolic blood pressure in WT and C3aR/C5aR1 double KO mice with no difference between the two groups as shown in figure 5A. Also area under the curve for systolic blood pressure showed no difference between WT and KO mice (figure 5A). An increased mortality was found in hypertensive mice without a significant difference between both genotypes (Figure 5B).

**Figure 5.**
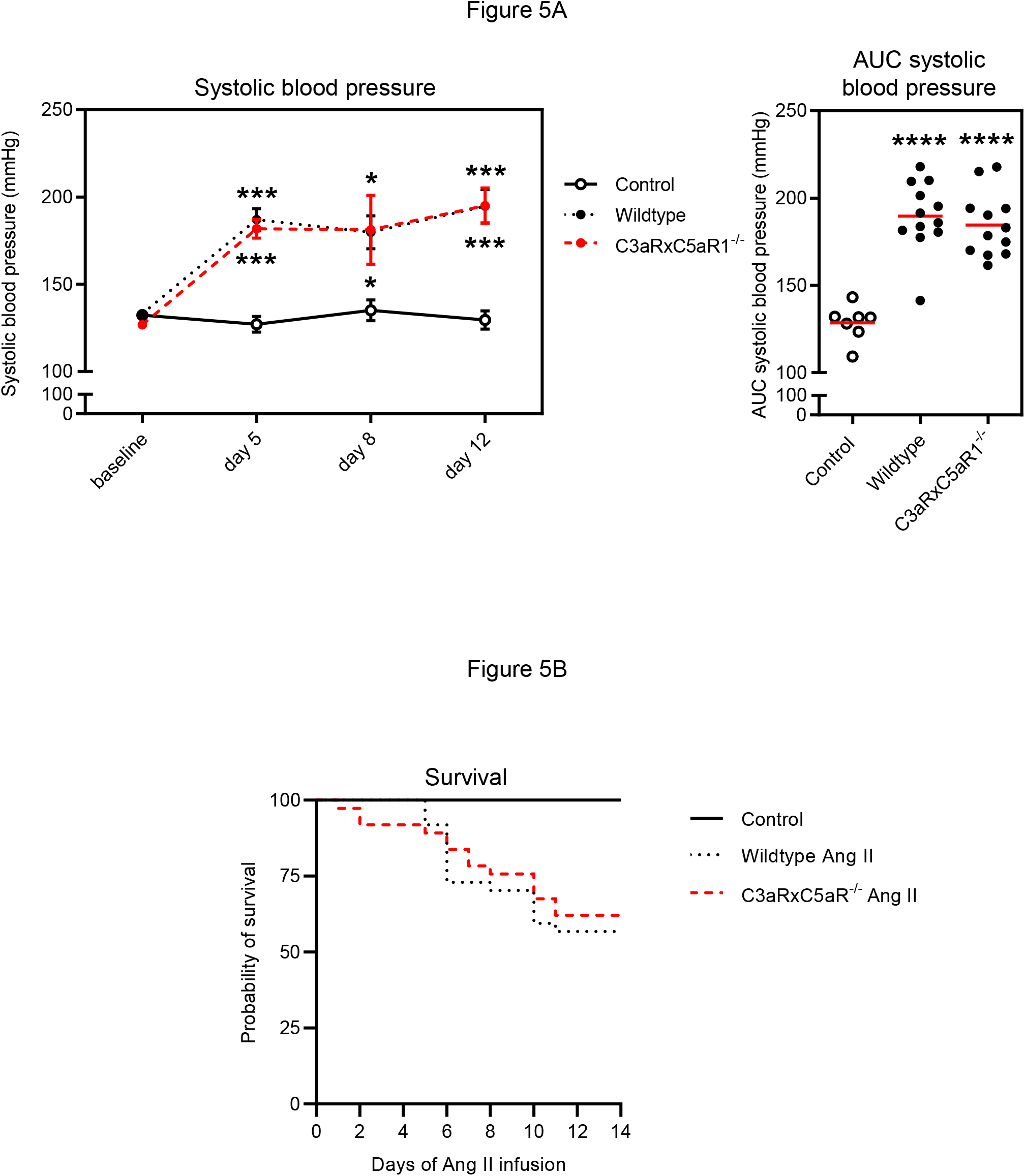
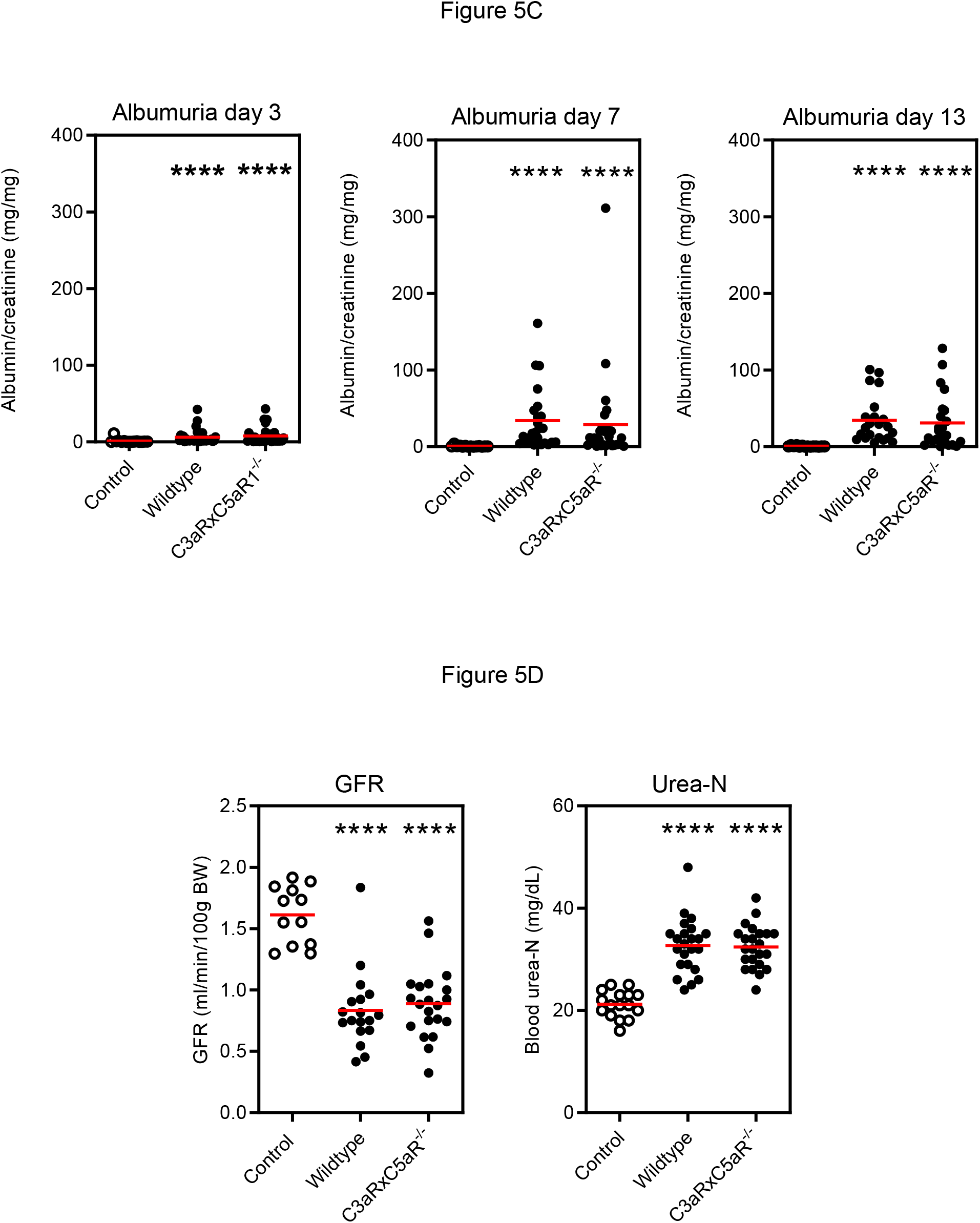
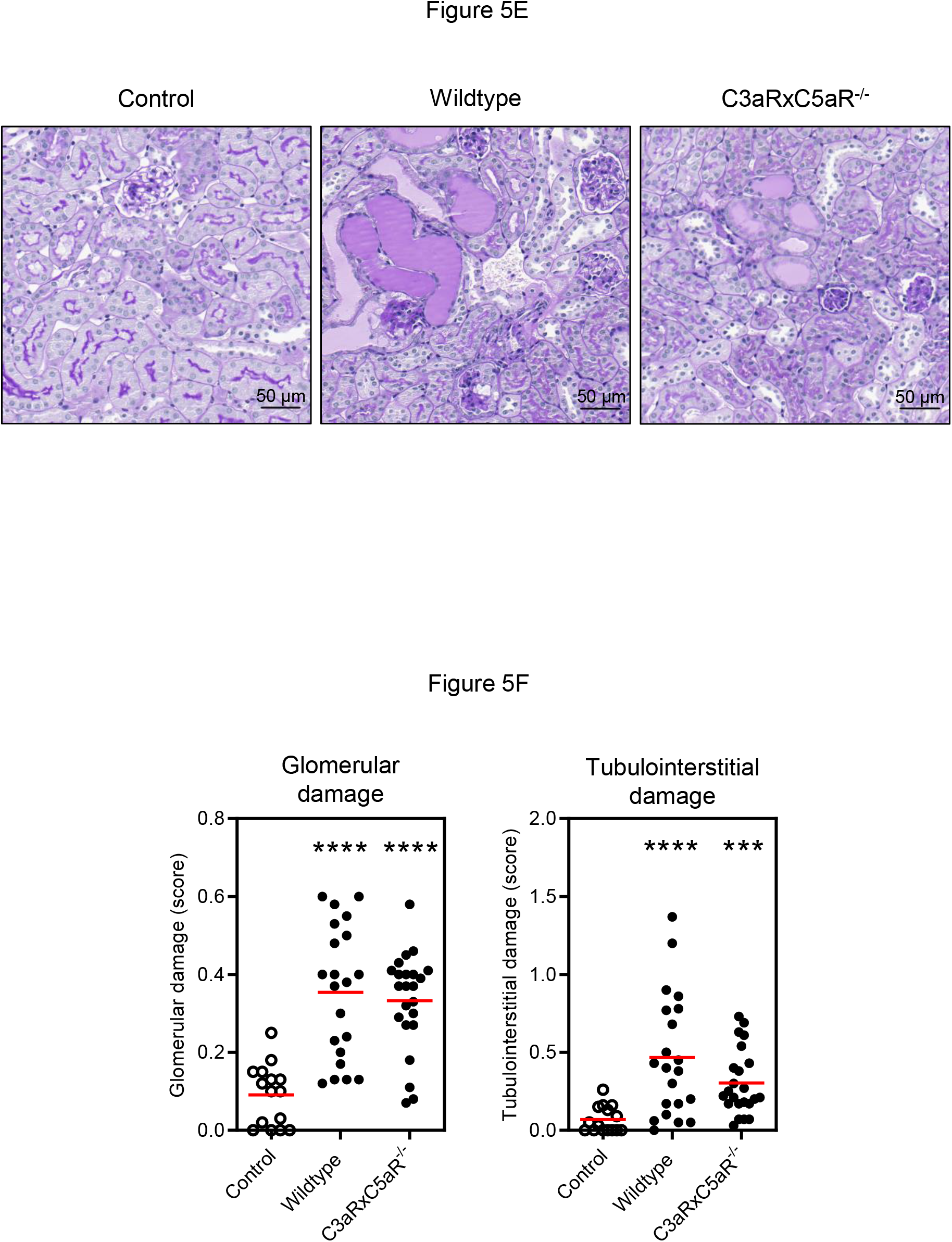
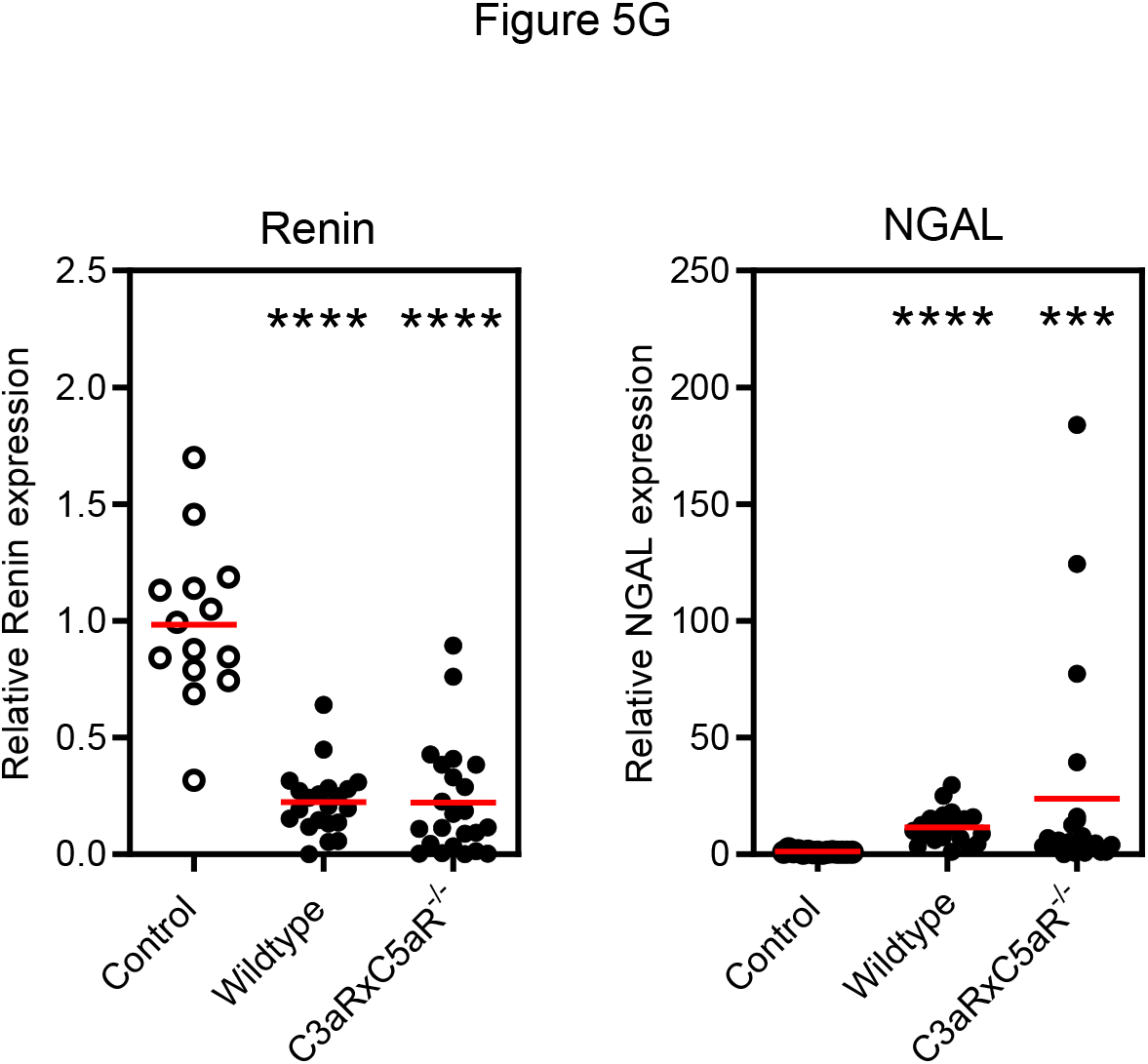
Systolic blood pressure and renal injury C3aR/C5aR1 double KO. A: Ang II infusion increased systolic blood pressure in WT and C3aR/C5aR1 double KO mice with no difference between the two groups. Shown over time and as area under the curve (AUC). B: No difference between WT und KO mice was found for mortality. C: Ang II induced heavy albuminuria without difference in both genotypes. D: Ang II decreased GFR in both groups. Accordingly, plasma values of urea-N were significantly elevated. E: PAS-stained sections revealed glomerular changes with mesangial expansion and sclerosis in both genotypes. F: Scoring of glomerular as well tubulointerstitial damage showed no significant differences between WT and C3aR/C5aR1-deficient mice. G: Suppression of renal renin and increased expression of NGAL was found in both hypertensive groups. White dots normotensive mice, black dots hypertensive mice. *=p<0.05, ***=p<0.001, ****=p<0.0001 vs. normotensive control

No albuminuria was found in C3aR/C5aR1^-/-^ deficient mice at the beginning of the experiment indicating that also the double KO mice do not have a basal renal phenotype (WT 0.39±0.03, C3aR/C5aR1^-/-^ 0.40±0.05 mg albumin/mg creatinine). After 3, 7 and 13 days of Ang II infusion heavy albuminuria developed in WT mice and KO mice but no significant difference was found between both groups (Figure 5C)

Next we measured GFR. A basal measurement before induction of hypertension was done. GFR averaged 1.56±0.23 in WT and 1.49±0.23 ml/min/100 g body weight in KO mice showing no difference in renal function before the start of the experiment. Ang II administration caused a drop of GFR in hypertensive mice on day 11 (52 % of controls in WT and 55% in KO mice) without difference between both genotypes (figure 5D). The drop in GFR was confirmed by increased plasma urea N values in both hypertensive groups (figure 5D). We next examined the kidney histology. Normal renal architecture was found in normotensive controls. Ang II infusion induced hypertensive glomerular injury with mesangial expansion and sclerosis in WT and C3aR/C5aR1^-/-^ mice shown in figure 5E. Semi-quantitative analysis of glomerular and tubulointerstitial injury (figure 5F) by scoring revealed similar changes in both genotypes.

The histology findings were supported by gene expression analysis. Suppression of renin was found in both hypertensive groups with no difference between both genotypes (Figure 5G). NGAL was upregulated and no difference was found (figure 5G).

Similar to the results in C3aR no major differences were found for myeloid cells and lymphocytes infiltrating the kidney as shown in suppl. figure 4A and B.

### Cardiac injury C3aR/C5aR1 double KO

No fibrosis was detected in normotensive mice and Ang II infusion induced extensive cardiac fibrosis as shown in figure 6A. However, scoring of the fibrosis revealed no difference between hypertensive WT and double KO mice (figure 6b). An increased relative heart weight was found in hypertensive mice but no difference between WT and KO mice was detected between both genotypes (Figure 6B). The presence of a significantly increased gene expression ANP and BNP mRNA in both genotypes further supports a similar cardiac injury in both genotypes (Figure 6C).

**Figure 6.**
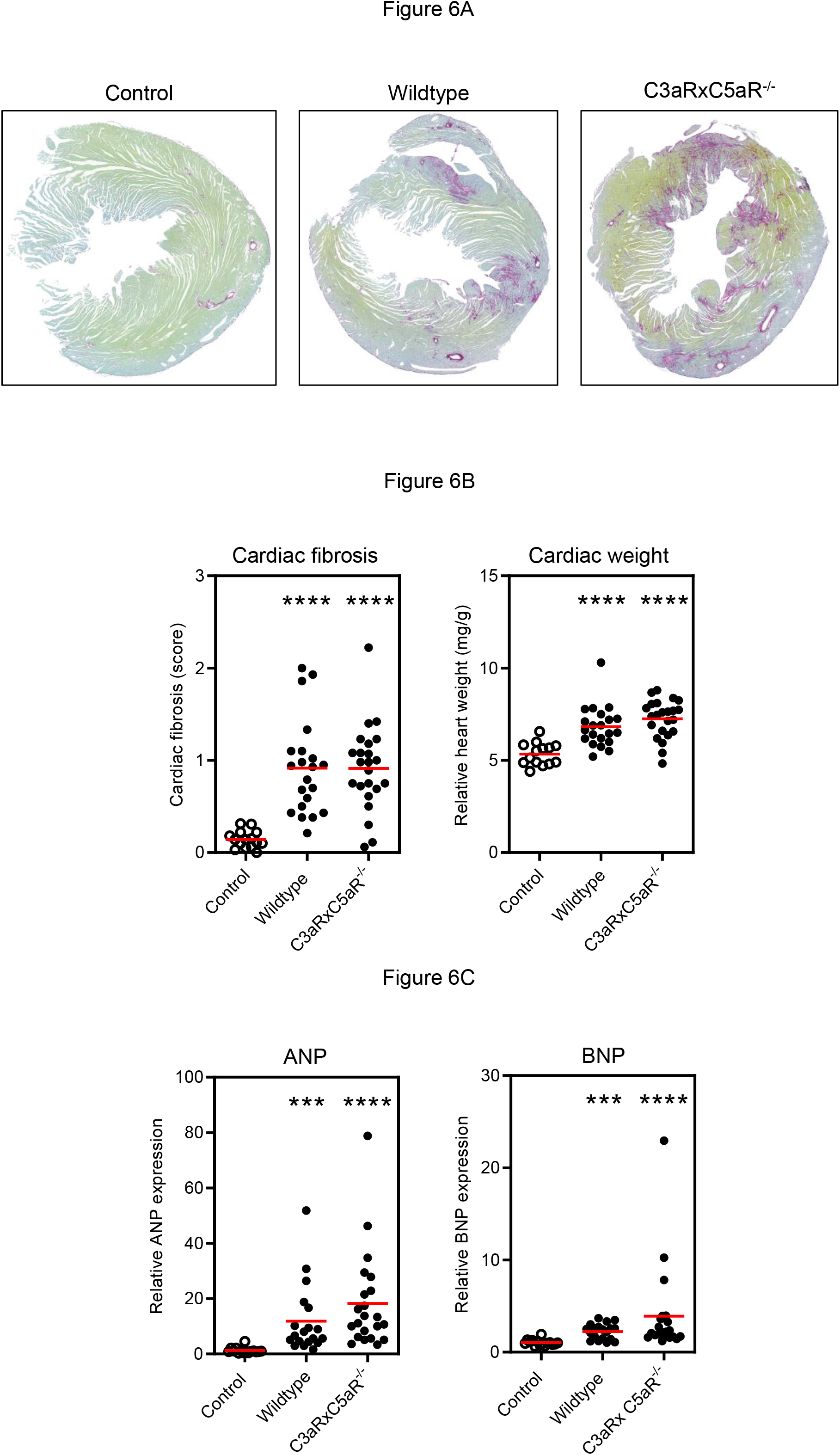
Cardiac injury C3aR/C5aR1 double KO. A: No fibrosis was seen in normotensive controls. Ang II-induced extensive cardiac fibrosis as shown in Sirius Red Fast Green stained cardiac sections. B: Cardiac fibrosis measured by scoring was not different between WT and KO mice. Relative heart weight was increased in hypertension with no difference between both groups. C: Hypertension induced increased cardiac expression of ANP and BNP and no difference was found between both genotypes. White dots normotensive mice, black dots hypertensive mice. ***=p<0.001, ****=p<0.0001 vs. control

### Tregs in hypertension

Renal infiltration of Tregs was quantified by flow cytometry analysis in a subset of mice. Moreover, gene expression of the transcription factor Foxp3 was also evaluated. Tregs can be divided into conventional Tregs (cTregs) and biTregs, which are double positive for Foxp3 and the IL-17 defining transcription factor RORγ. cTregs and biTregs have at least to our knowledge not been evaluated in hypertension. Representative FACS blot of Tregs as well as cTregs and biTregs are shown in figure 7A and B. Quantification is shown figure 7C. Ang II induced a small increase of Tregs in WT mice. A significant increased number of Tregs was found in the in the C3aRxC5aR1 double KO mice compared to normotensive controls. However, no significant difference was found between hypertensive WT and KO mice. Treg subtypes followed this pattern. FACS analysis quantifies the number of positive cells. Mean fluorescent intensity (MFI) quantifies additionally the expression level in each cell and gives a better picture. However, as shown in figure 7D, MFI data of Foxp3 were not different the pattern found for positive cells.

**Figure 7.**
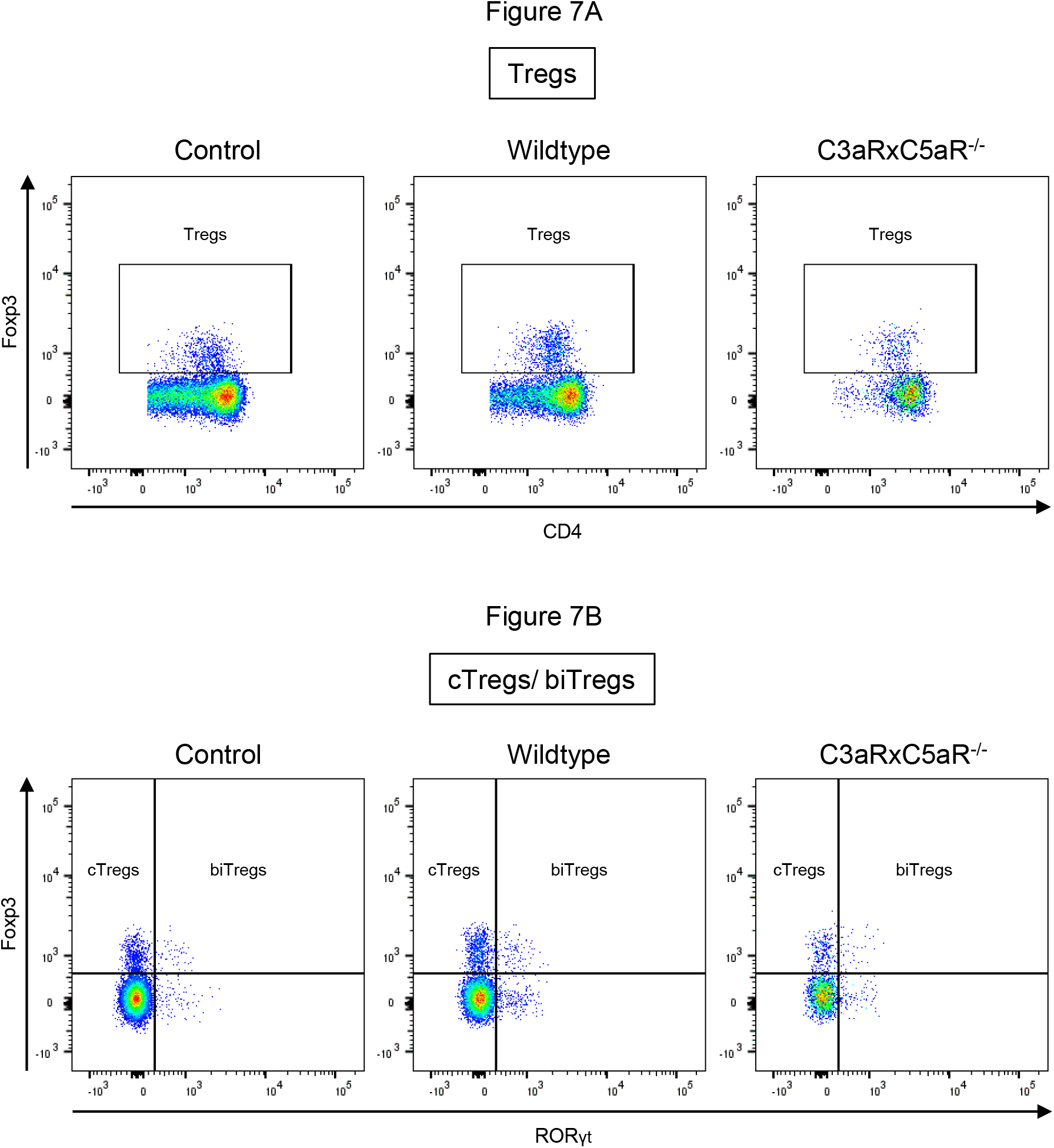
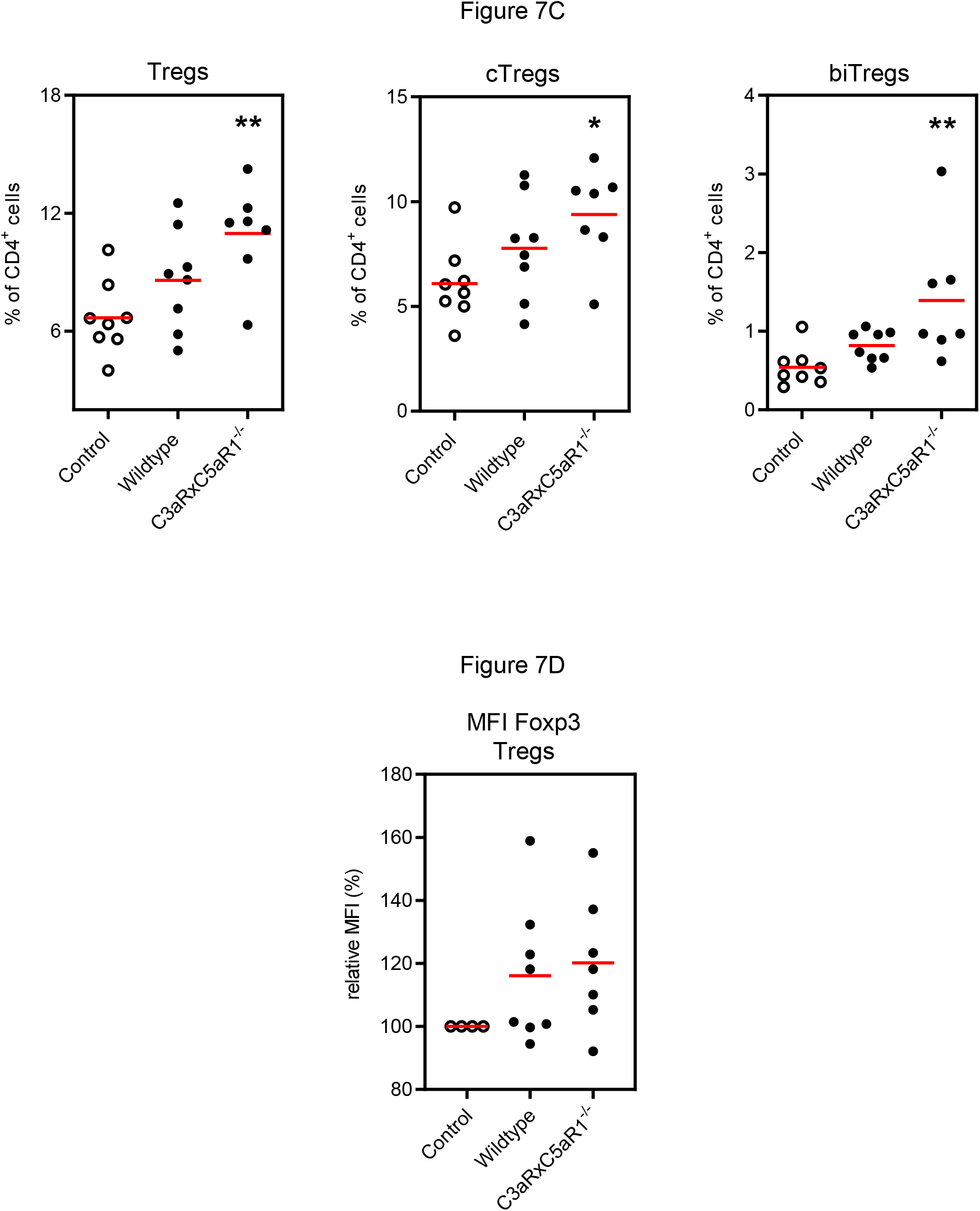
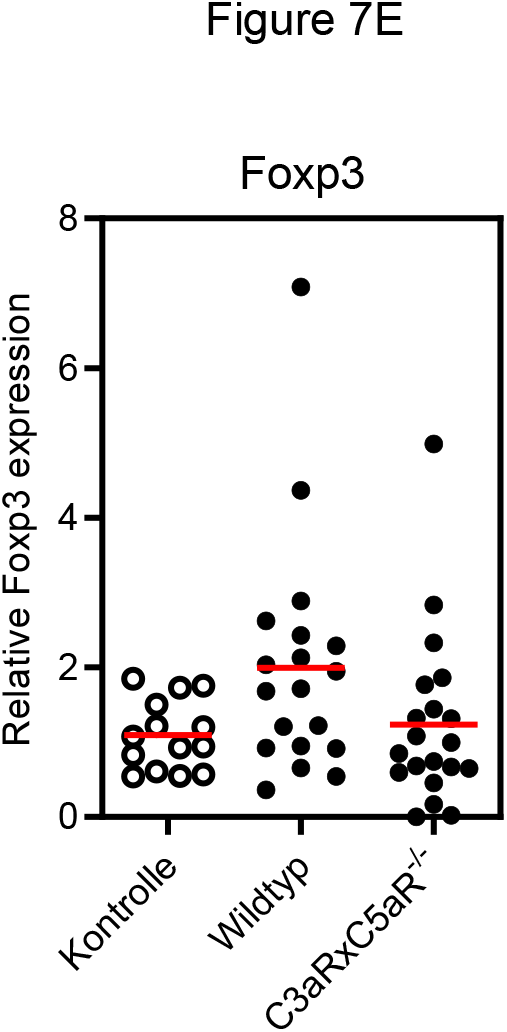
Tregs in experimental hypertension. A: Representative FACS blot of Tregs in controls, hypertensive WT and C3R/C5aR1- deficient mice are shown. B: FACS blots gated for cTregs and biTregs. C: Quantification of Tregs, cTregs and biTregs. D: Mean fluorescent intensity (MFI) of Foxp3 in Tregs. E: Renal gene expression of Foxp3. White dots normotensive mice, black dots hypertensive mice. **=p<0.01 vs. control

We also examined gene expression of the Treg defining transcription factor Foxp3 in renal tissue. Ang II induced a small increase in WT and double KO mice without difference between both genotypes (figure 7E).

To examine whether the Treg pattern found in the present study is specific for Ang II infusion or can also be found in other models of hypertension we determined the number of Tregs infiltrating the kidney in DOCA salt hypertension. 6 weeks of DOCA salt treatment resulted in a mild increase of Tregs infiltrating the kidney as shown in suppl. figure 5A. The increase in biTregs was significant. This clearly shows that in different models of hypertension Tregs are rather upregulated than downregulated in the kidney. The presented experiments were performed in Balb/c mice. Since many hypertension studies are performed in mice of the C57black strain, we also studied the number of Tregs infiltrating the kidney in C57black mice to see whether the Treg pattern in hypertension is strain specific. As shown in suppl. figure 5B Ang II as well as DOCA salt resulted in a significant increased number of Tregs as well as c and biTregs infiltrating the kidney. The increase was more pronounced than in mice of the Balb/c strain.

Finally, we isolated Tregs from a Foxp3 reporter mouse allowing isolation of very clean Treg population (>99.9%) to examine presence of C3aR and C5aR1 especially on Tregs. Gene expression by realtime PCR showed low expression of C3aR (ct 33.77±0.60) whereas C5aR1 was extremely low (ct 37.01±3.00). Cts of 30-37 are positive reactions indicative of moderate amounts of target nucleic acid. Cts of 38-40 are weak reactions indicative of minimal amounts of target nucleic acid which could represent a contaminant. This confirms that Tregs express moderately C3aR but not C5aR1.

## Discussion

The innate and adaptive immune system are increasingly recognized to play a decisive role in arterial hypertension and hypertension-induced end organ damage (Wenzel *et al*., 2016). The complement system is an ancient part of innate immunity. Recent data suggest that the complement system also contributes to arterial hypertension and hypertensive end-organ damage (Wenzel *et al*., 2021c). The current study clarifies the role of the anaphylatoxin receptor C3a and C5aR1 and the role of Tregs in arterial hypertension and hypertensive end-organ damage.

On the basis of their cytokine production profile and expression of specific transcription factors, CD4^+^ T cells are classified into four major subsets: TH1, TH2, TH17, and Tregs. The main function of Tregs is the maintenance of immunological tolerance, i.e., by producing IL-10. Adoptive transfer of Tregs lowers blood pressure and ameliorates cardiac and renal injury in different models of hypertension. Moreover, IL-2/anti-IL-2 immune complex treatment of hypertensive mice expands Tregs and reduces aortic stiffening as summarized recently (Wenzel *et al*., 2021b). Tregs can be divided into conventional Tregs and so called biTregs or Tregs 17 cells that co-express the Treg characteristic transcription factor Foxp3 together with RORγt, one of the defining transcription factors of the Th17 cells (Herrnstadt *et al*., 2021; Kluger *et al*., 2016; Lochner *et al*., 2008). Abundance of cTregs and biTregs have not been examined in hypertension yet.

Complement activation produces cleavage fragments like the anaphylatoxin C3a and C5a. C3a and C5a are proinflammatory effector molecule and exerts pleiotropic immunoregulatory functions, most of which are mediated through cognate interactions with the G protein-coupled complement receptors C3aR and C5aR1. The role of C3a and the C3aR in hypertension has not been examined in detail. In C3aR-deficient mice, Zhang et al. observed a trend for increased cardiac injury when compared with wildtype mice after Ang II infusion (Zhang *et al*., 2014a).

More data are available on the C5a-C5aR1 axis. Significantly elevated C5a serum levels were observed in hypertension (Zhang *et al*., 2014a). In line with this, infusion of Ang II causing arterial hypertension led to increased systemic anaphylatoxin generation in mice (Zhang *et al*., 2014a). Conversely, C5aR1-deficient mice exhibited markedly reduced cardiac remodeling and inflammation after Ang II infusion. Similarly, pharmacological inhibition of C5a production by an anti-C5 monoclonal antibody or C5aR1 targeting with an inhibitor (PMX53) reproduced the effects of C5aR1 deficiency (Zhang *et al*., 2014a; Zhang *et al*., 2014b). We also found a blood pressure-independent nephroprotective effect of C5aR1 deficiency in our accelerated model of hypertension in mice (Weiss *et al*., 2016).

Interestingly, the anaphylatoxins and their receptors are known as regulators of Treg activity: C3a/C3aR and C5a/C5aR1 inhibit the function of CD4^+^ Foxp3^+^ circulating Tregs and absence of C3aR and C5aR1 signals in CD4^+^ T cells induces spontaneous Foxp3^+^ Treg differentiation upon T cells activation (Kwan *et al*., 2013; Strainic *et al*., 2013).

Despite the large number of clinical trials targeting C5, C5a and C5aR1 trials targeting C3, C3a and C3aR are less. Protective effects of C3aR deficiency or antagonism have been reported in membranous nephropathy, diabetic nephropathy, focal segmental glomerulosclerosis, ischemia reperfusion injury and other renal diseases (Morigi *et al*., 2020, Gao *et al*., 2022 and review in Gao *et al*., 2020). However, in the present study, C3aR deficiency clearly did not protect the kidney from hypertensive injury. More in-depth investigations and awareness of the role of C3a-C3aR in kidney disease are needed to clarify whether C3aR inhibition has a translational potential for treatment of chronic kidney disease in humans.

In this context Chen et al. suggested an interesting approach to lower blood pressure and decrease hypertensive injury. In a series of very elegant experiments they showed that Ang II-induced hypertension is characterized by a decreased number of Tregs in the kidney. Moreover, they found that C3aR and C5aR1 double deficiency decreased blood pressure in response to Ang II, compared with wildtype mice, via activating Foxp3^+^ Tregs (Chen *et al*., 2018).

We first were interested in the expression of the anaphylatoxin receptors and Foxp3 in human hypertensive nephropathy. For this we first examined C3aR, C5aR1 and Foxp3 expression in the European Renal cDNA biopsy bank and found a significantly upregulated gene expression of C3aR and Foxp3. To further pin down the cellular source of this expression, we examined available kidney tissue data. Single cell data analysis from a kidney tissue atlas revealed that C3aR and C5aR1 are almost exclusively expressed in myeloid cells. Interestingly both receptors were upregulated in hypertension. Gene expression of regulatory T cell main transcription factor Foxp3 was found in parts of the T cell cluster, showing overlap expression with C3aR but not for C5aR1.

We next turned our attention to C3aR and C5aR1 in Ang II induced hypertension in mice. We examined hypertension as well as renal and cardiac hypertensive injury in great detail and neither C3aR- nor C3aR/C5aR1 double deficiency ameliorated hypertension or renal and cardiac injury.

We also examined the number of Tregs in mice with Ang II infusion and the effect of a double KO of C3aR and C5aR1. By using gene expression and flow cytometry we found no downregulation of Tregs in the kidney of Ang II infused mice but rather a mild upregulation. Moreover, hypertensive C3aRxC5aR1 double KO mice did not show a significant higher number of Tregs compared to hypertensive WT mice. This was also valid for cTregs and biTregs. Using a tdTomato-C3aR knock-in mouse we found very low expression of C3aR in lymphocytes confirming previously published data (Quell *et al*., 2017). In addition we also were unable to find C5aR1 expression in lymphocytes isolated from the kidney in our previous work (Weiss *et al*., 2016, Karsten *et al*., 2015). In contrast, Chen et al. found increased expression of C3aR and C5aR1 on Tregs isolated from the kidney using FACS analysis. The reason for this discrepancy is unknown but may be caused by the different techniques used. Antibody based quantification of G protein receptors as done by Chen et al. is prone to false positive results whereas reporter based approaches have a high specificity. To further analyse expression of both receptors in Tregs, we isolated Tregs using a Foxp3 reporter mouse. Realtime PCR analysis revealed low expression of C3aR but did not detect C5aR1. Therefore, the murine data confirm the human single cell data.

To examine whether the renal Treg pattern found in the present study is specific for Ang II infusion or can also be found in other models of hypertension we also studied renal Tregs in DOCA salt hypertension. Also in this model renal Tregs were rather up- than downregulated.

It should be mentioned that the present study was done in mice of the Balb/c strain whereas in the study of Chen et al. C57black mice were used. However, Ang II infusion as well as DOCA salt hypertension in C57black mice resulted in an increased number of Tregs in the kidney.

It is difficult to study hypertensive renal injury in mice of the C57black or Balb/c strain. Due to the effect that these mice are resistant to hypertensive renal injury by Ang II infusion (Wesseling *et al*., 2005), we induced an accelerated model of hypertension by combining Ang II infusion with unilateral nephrectomy and application of salt in the drinking water. Ang II infusion for 2 weeks does not induce noteworthy renal injury. Chen et al. found increased glomerular hypertrophy and interstitial collagen deposition examined by picrosirius red staining. Unilateral nephrectomy, salt and Ang II in our model of accelerated hypertension induce severe albuminuria, glomerular and tubulointerstitial injury as well as s significant drop in GFR.

Barhoumi et al. also described a decreased number of renal Tregs evaluated by immunohistochemistry in Ang II infused mice (Barhoumi *et al*., 2011). In aldosterone + salt induced hypertension the same group found a nonsignificant tendency for a reduced number of Tregs in renal cortex (Kasal *et al*., 2012). The reason for the difference remains unclear.

### Perspectives

Our data underscore the complexity of the immune and complement system in hypertensive end-organ damage. C3aR and C5aR1 expression is almost exclusively found in myeloid cells in the human kidney. Expression is increased in hypertensive nephropathy in humans. Hypertension in mice and men is not a characterized by a decreased number of Tregs. KO of the anaphylatoxin receptor C3aR alone or in combination with C5aR1 does not upregulate significantly infiltration of Tregs into the kidney and does not lower blood pressure and hypertensive end organ damage in Ang II induced hypertension.

In contrast to the work by Chen et al. our study does not suggest that targeting the anaphylatoxin receptors C3aR alone or in combination with C5aR1 is useful to treat cardiovascular disease in hypertensive patients.

## Data Availability

All data produced in the present study are available upon reasonable request to the authors

## Acknowledgement

We thank S. Gatzemeier and Sonja Wulf for excellent technical assistance.

---

**Supplement figure 1.**
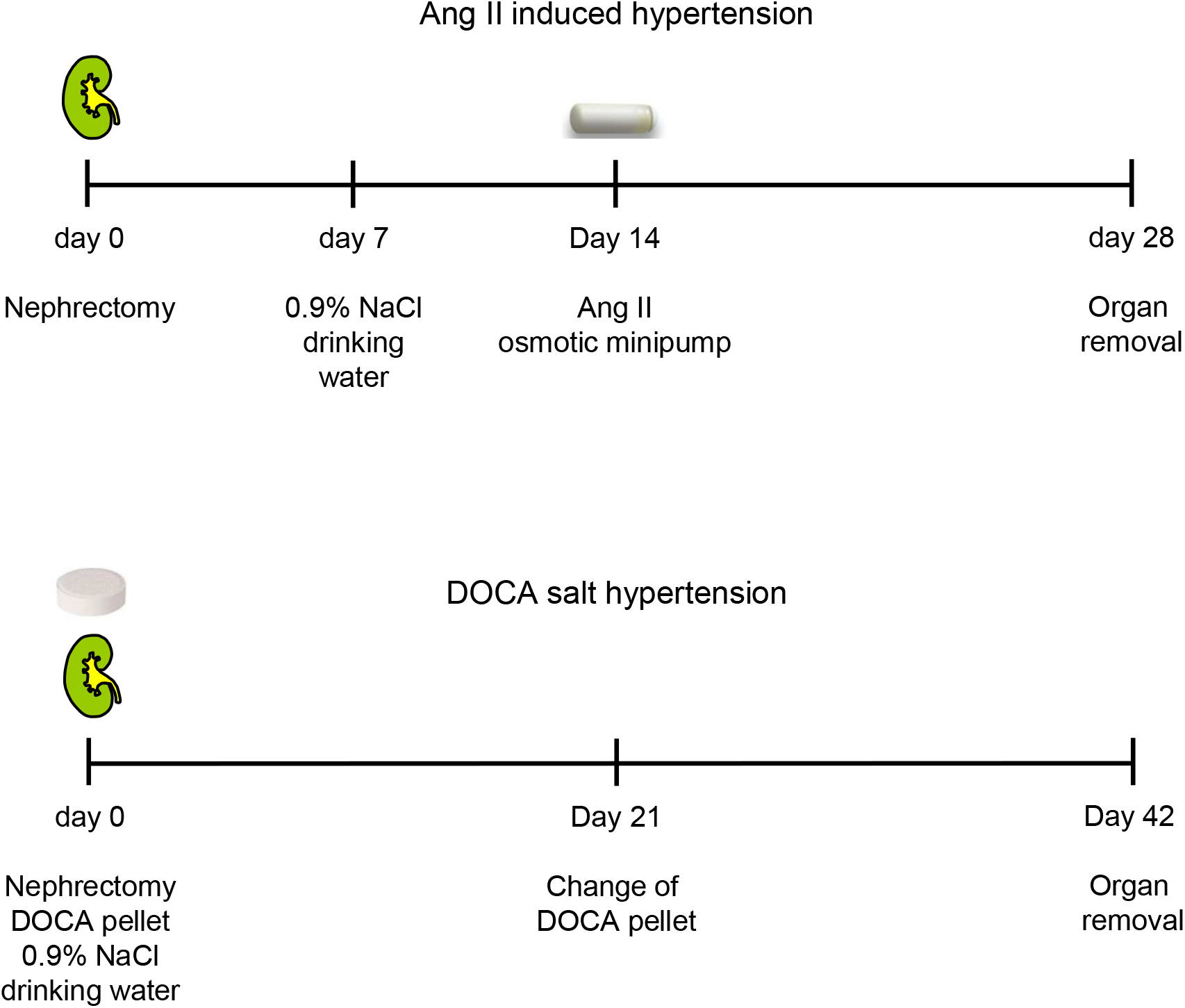
Ang II and DOCA salt induced hypertension. Time course of the accelerated model of Ang II as well as DOCA salt induced hypertension.

**Supplement figure 2.**
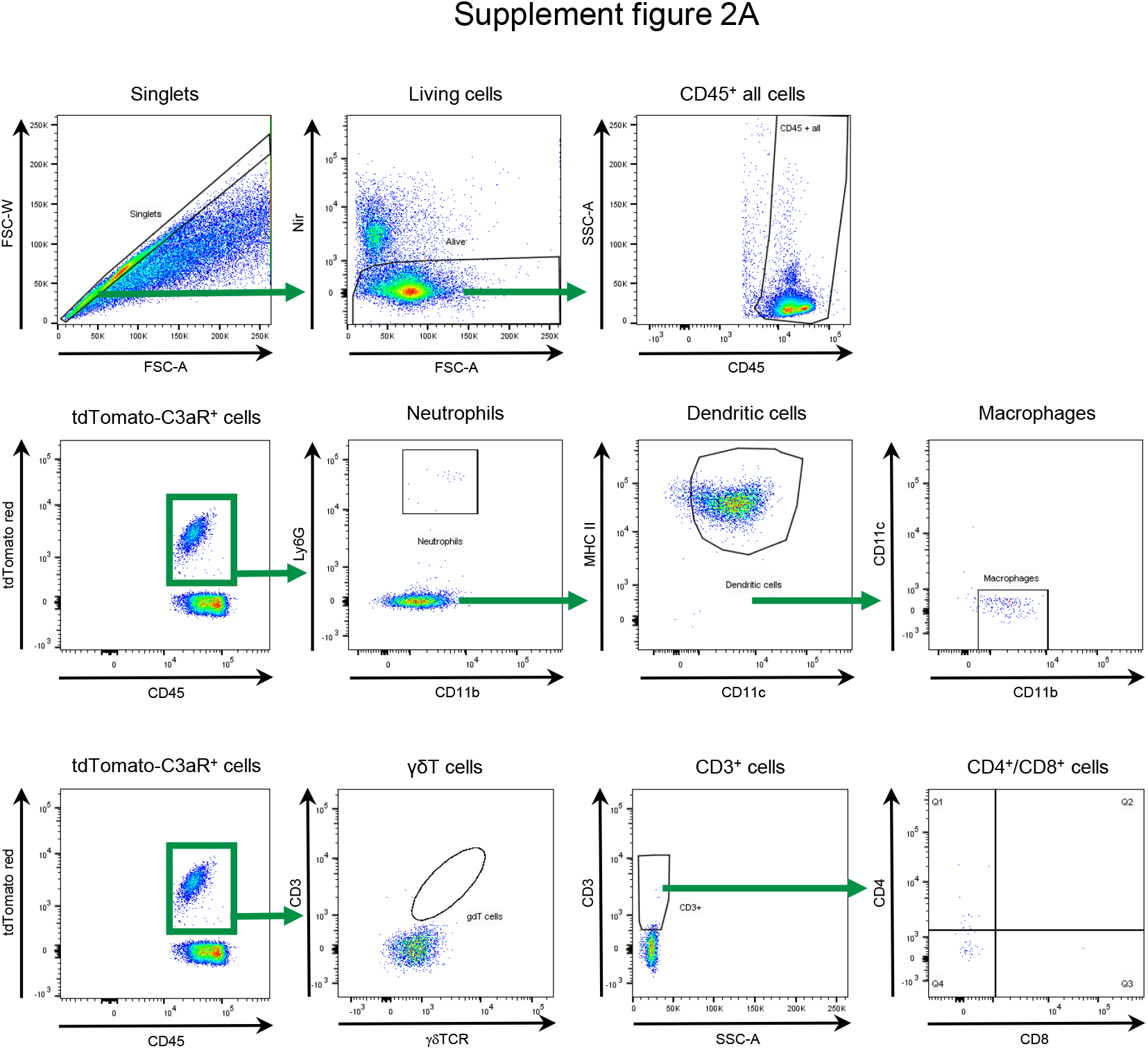
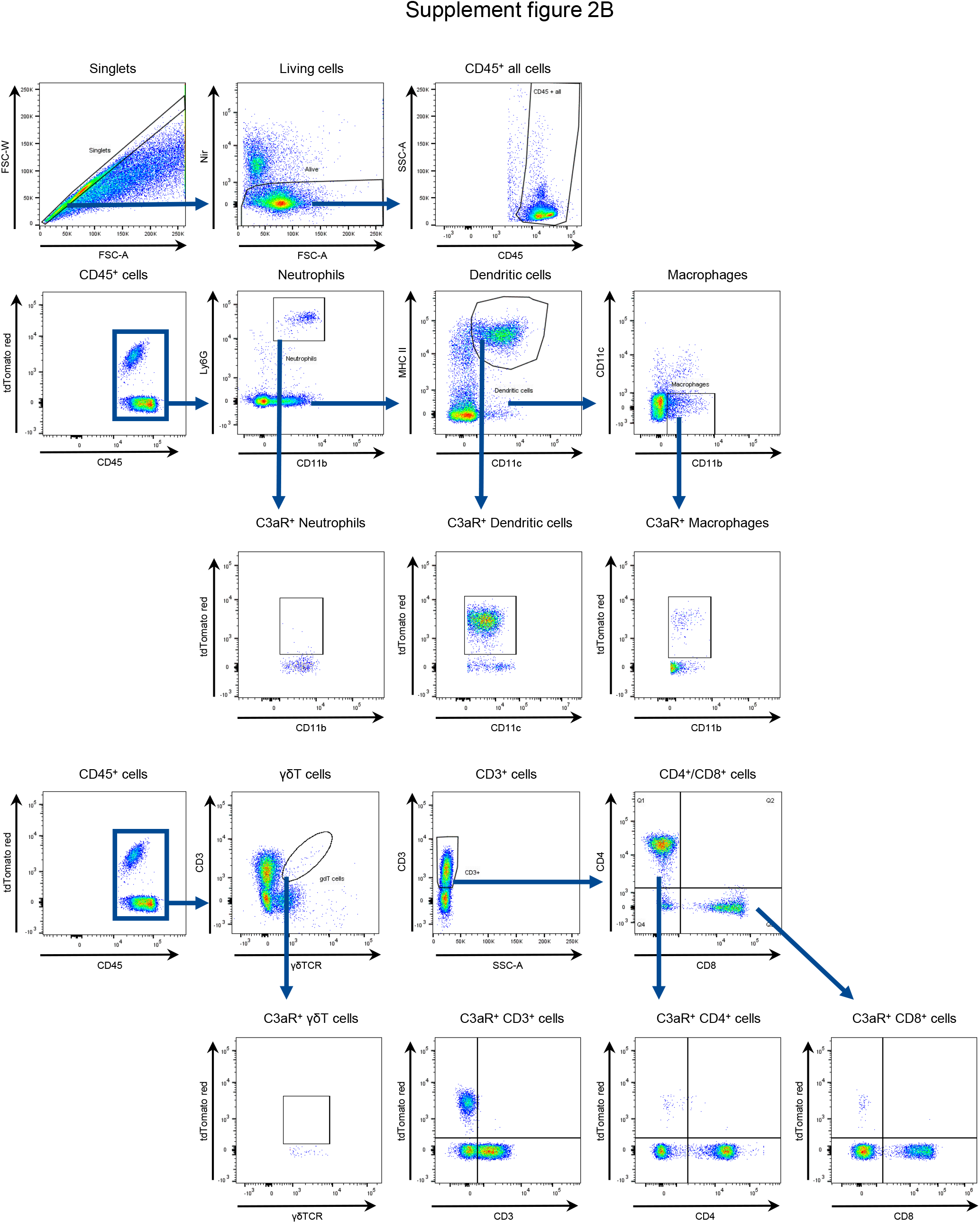
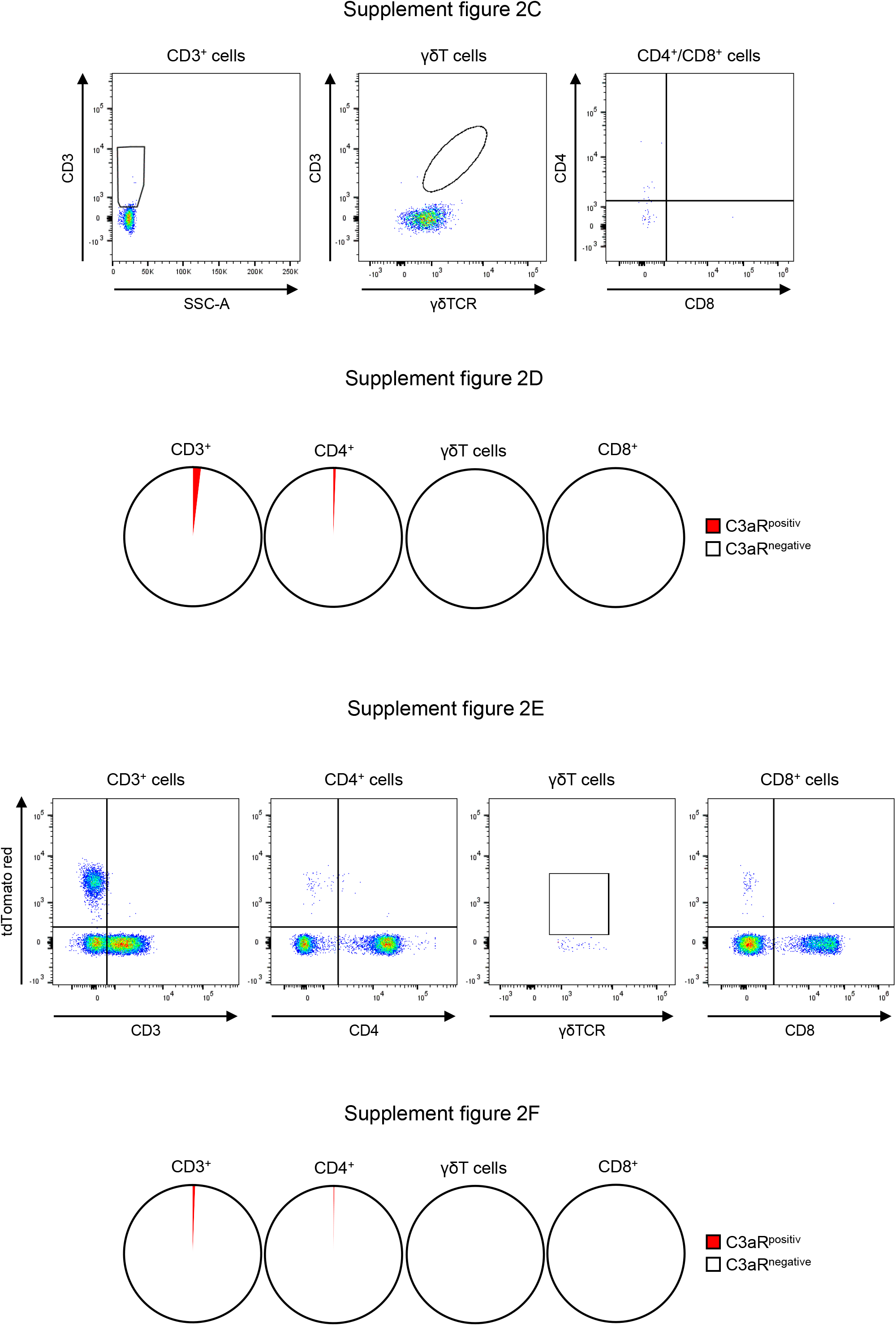
**A Identification of C3aR-positive cell populations in the kidney** Gating strategy: Living single cells were gated by forward scatter width (FSC-W) and forward scatter area (FSC-A) and by Nir. The C3aR-positive cells were identified by a tdTomato red and CD45 positive signal. Afterwards Neutrophils were determined by Ly6G and CD11b expression. The Dendritic cells were identified by MHC II and CD11c staining. Macrophages were positive for CD11b but negative for CD11c. The whole T cell population was distinguished by SSC-A and CD3 expression. γδ T cells were identified by CD3 and γδ T cell receptor (γδTCR) expression. CD3 positive T cells were further analyzed for their expression of CD4 and CD8 to determine these subpopulations. **Supplement figure 2B to 2F Identification of C3aR-positive cells in the kidney 2B** Gating strategy: Living single cells were gated by forward scatter width (FSC-W) and forward scatter area (FSC-A) and by Nir. The leukocytes were identified via CD45 surface staining. Afterwards Neutrophils were determined by Ly6G and CD11b expression. The Dendritic cells were identified by MHC II and CD11c staining. Macrophages were positive for CD11b but negative for CD11c. The whole T cell population was distinguished by SSC-A and CD3 expression. γδ T cells were identified by CD3 and γδ T cell receptor (γδTCR) expression. CD3 positive T cells were further analyzed for their expression of CD4 and CD8 to determine these subpopulations. The C3aR-positive cells were determined after identifying the leukocyte and lymphocyte subpopulations. **2C** Plots and **2D** diagrams: 1,9% of the C3aR-positive leukocytes are CD3^+^ and 0,6% are CD4^+^ lymphocytes. **2E** Plots and **2F** diagrams: 0,6% of all CD3^+^ and 0,2% all CD4^+^ lymphocytes are positive for the C3aR-positive signal.

**Supplement figure 3.**
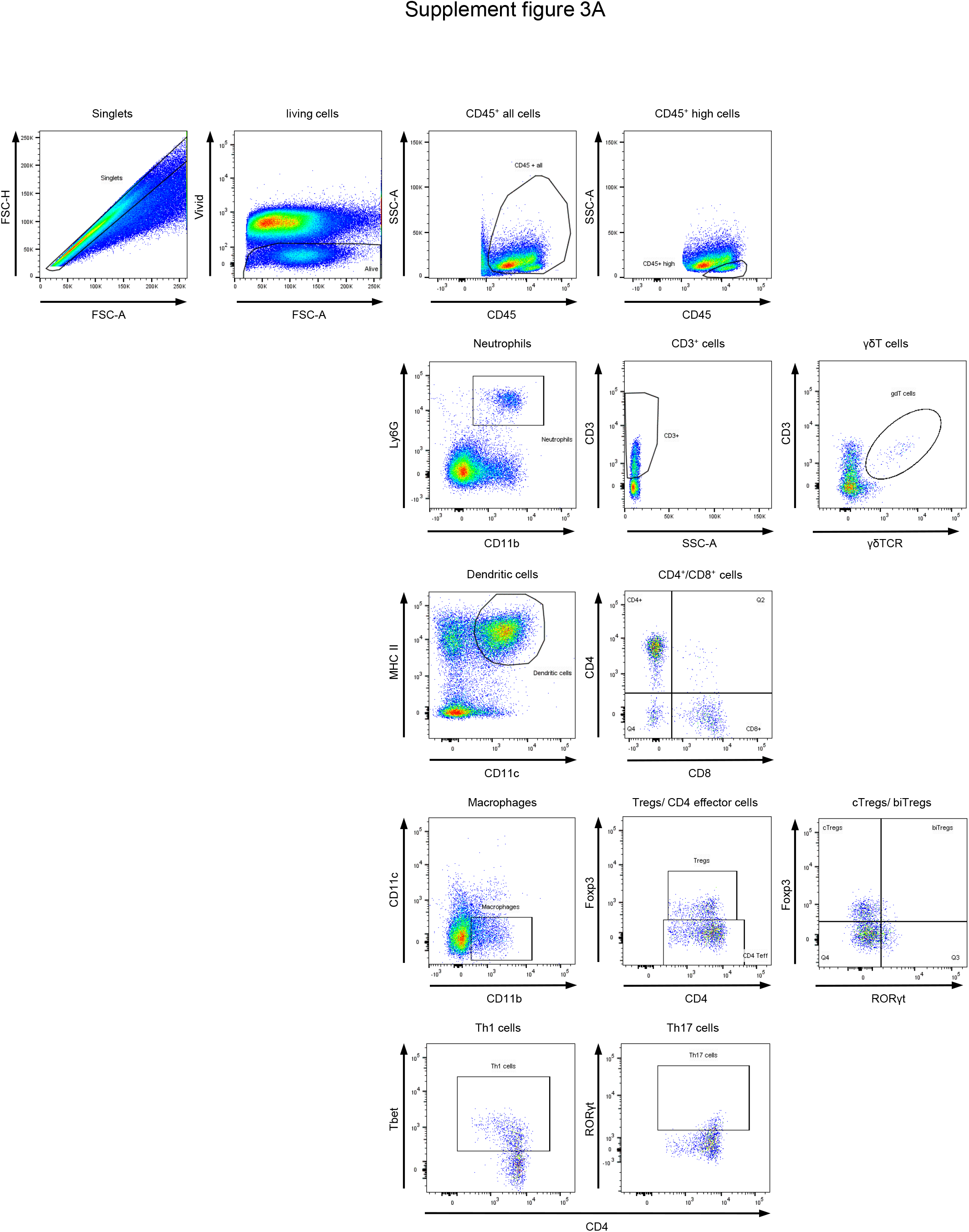
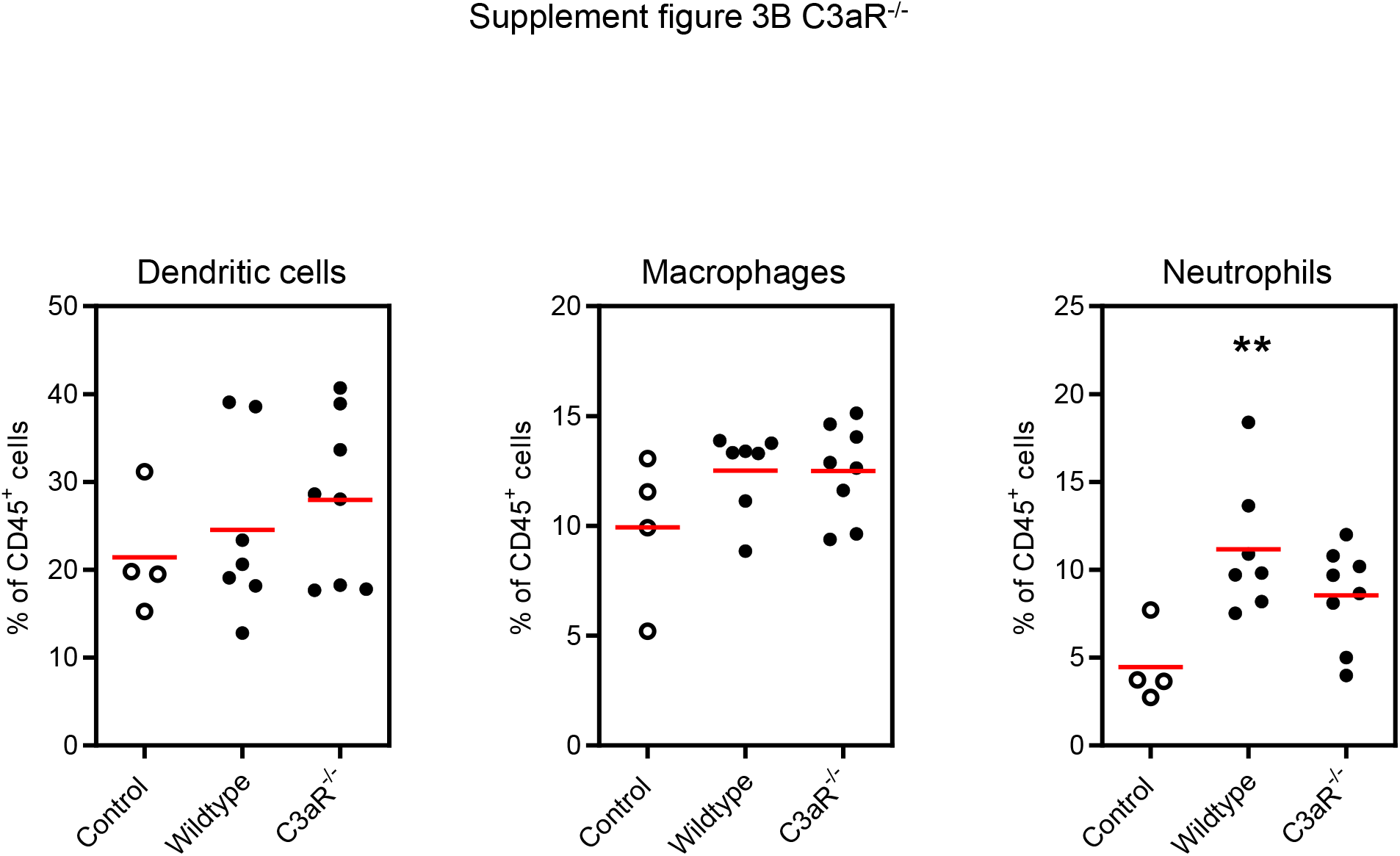
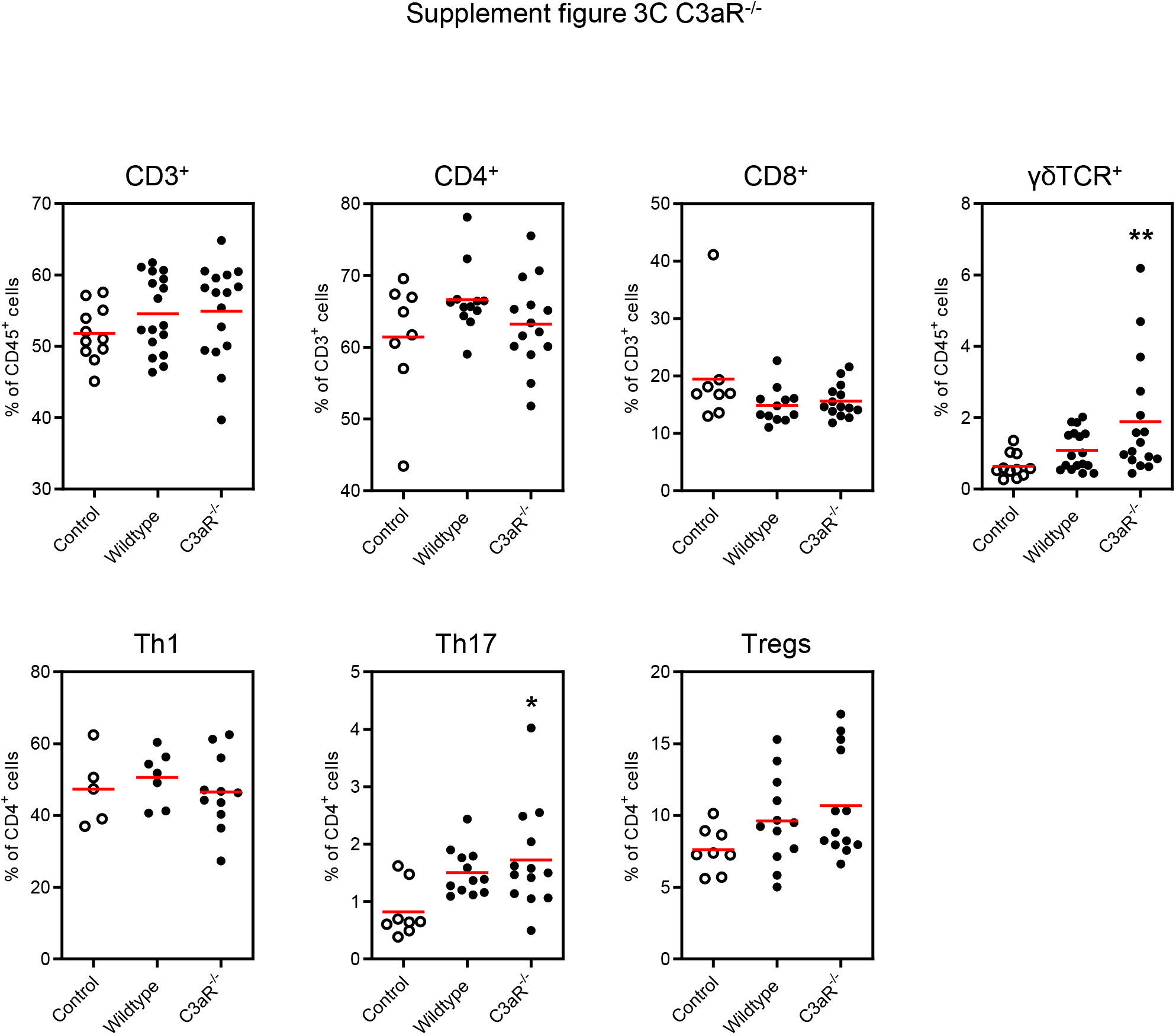
**A Infiltration of leukocytes into the kidney** A: Gating strategy: Leukocytes were identified via CD45 surface staining after gating for living single cells by forward scatter width (FSC-W) and forward scatter area (FSC-A) and by Vivid. Neutrophils were determined by Ly6G and CD11b expression. For determination of dendritic cells and macrophages, neutrophils were excluded. Dendritic cells were identified by MHC II and CD11c staining. Macrophages were positive for CD11b but negative for CD11c. Lymphocytes were determined by SSC-A and CD45^high^ positive expression. The whole T cell population was distinguished by SSC-A and CD3 expression. γδ T cells were identified by CD3 and γδ T cell receptor (γδTCR) expression. CD3 positive T cells were further analyzed for their expression of CD4 to determine CD4 positive T helper cells. CD4 positive T helper cells were identified as FoxP3 negative and either Tbet positive (TH1 cells) or RORγt positive (TH17 cells). **B Infiltration of myeloid cells into the kidney** The frequencies of dendritic cells, macrophages and neutrophils (PMNs) were modestly increased and similar in hypertensive wildtype and knockout mice. White dots normotensive mice, black dots hypertensive mice. **=p<0.01 vs. control **C Infiltration of lymphocytes into the kidney** The frequencies of CD3, CD4, CD8 and γδ T cells were not different between both genotypes in hypertension. The same was valid for the T cell subtypes TH1, TH17 and Tregs. White dots normotensive mice, black dots hypertensive mice. *p<0.05, **=p<0.001 vs. control.

**Supplement figure 4.**
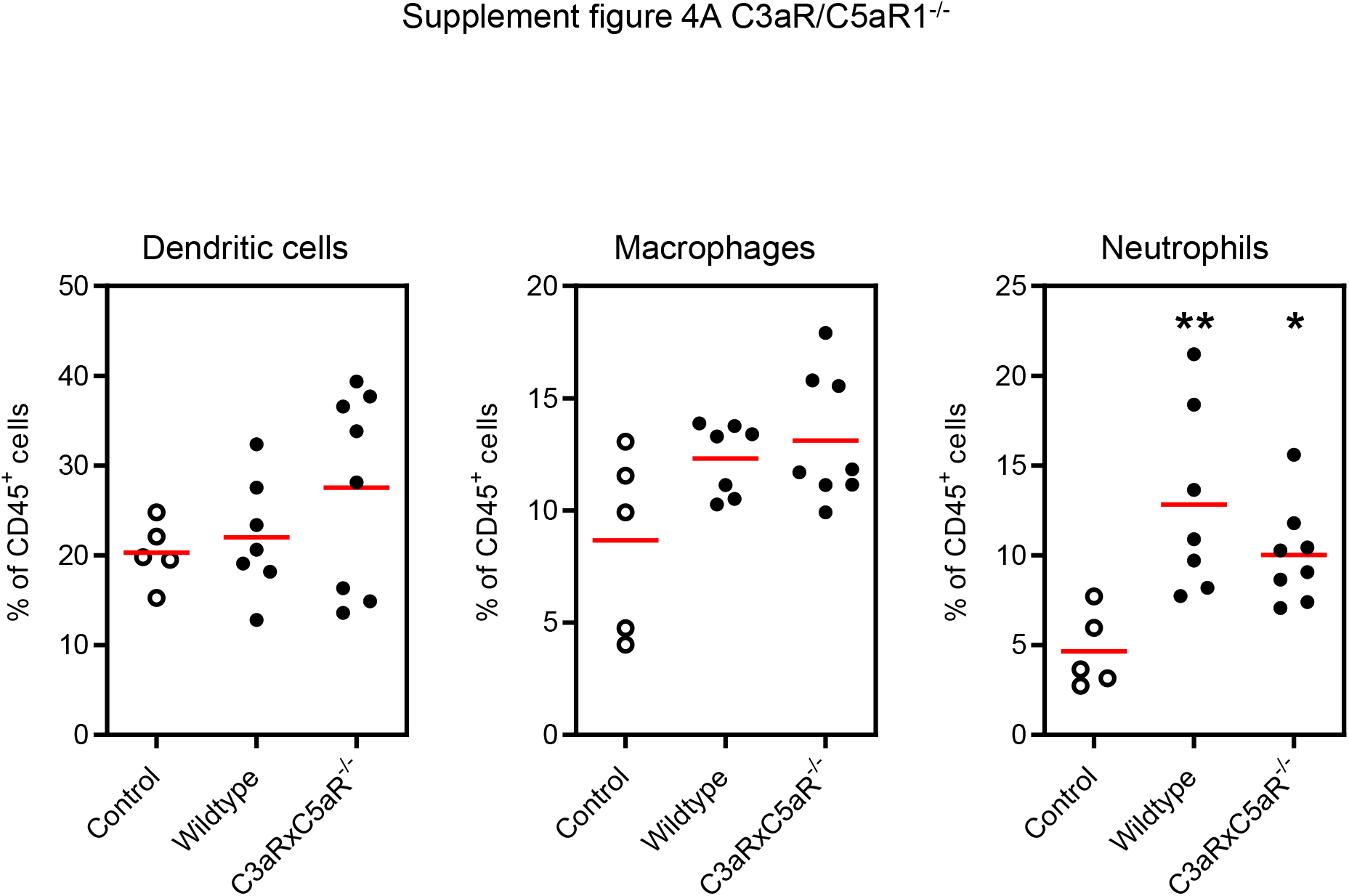
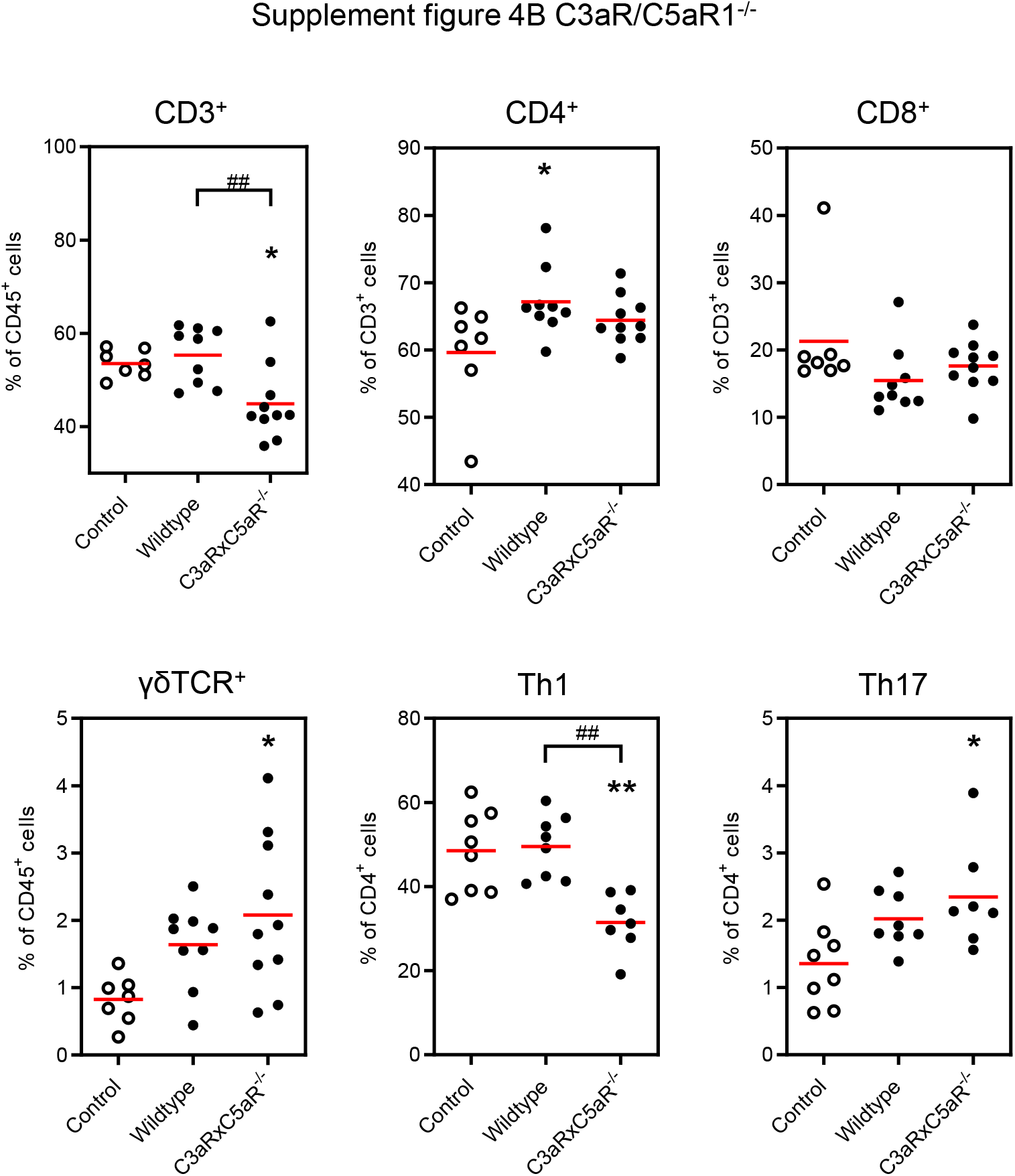
**A Infiltration of myeloid cells into the kidney** The frequencies of dendritic cells, macrophages and neutrophils were modestly increased and similar in hypertensive wildtype and knockout mice. White dots normotensive mice, black dots hypertensive mice. *p<0.05, **=p<0.001 vs. control. **B Infiltration of lymphocytes into the kidney** A lower number of CD3^+^ cells was found in hypertensive C3aR/C5aR1^-/-^ mice compared to normotensive controls and hypertensive WT mice. The frequencies of CD4, CD8 and γδ T cells were not different between both genotypes in hypertension. The same was valid for the T cell subtypes TH1 and TH17. White dots normotensive mice, black dots hypertensive mice. *p<0.05 vs. control.

**Supplement figure 5.**
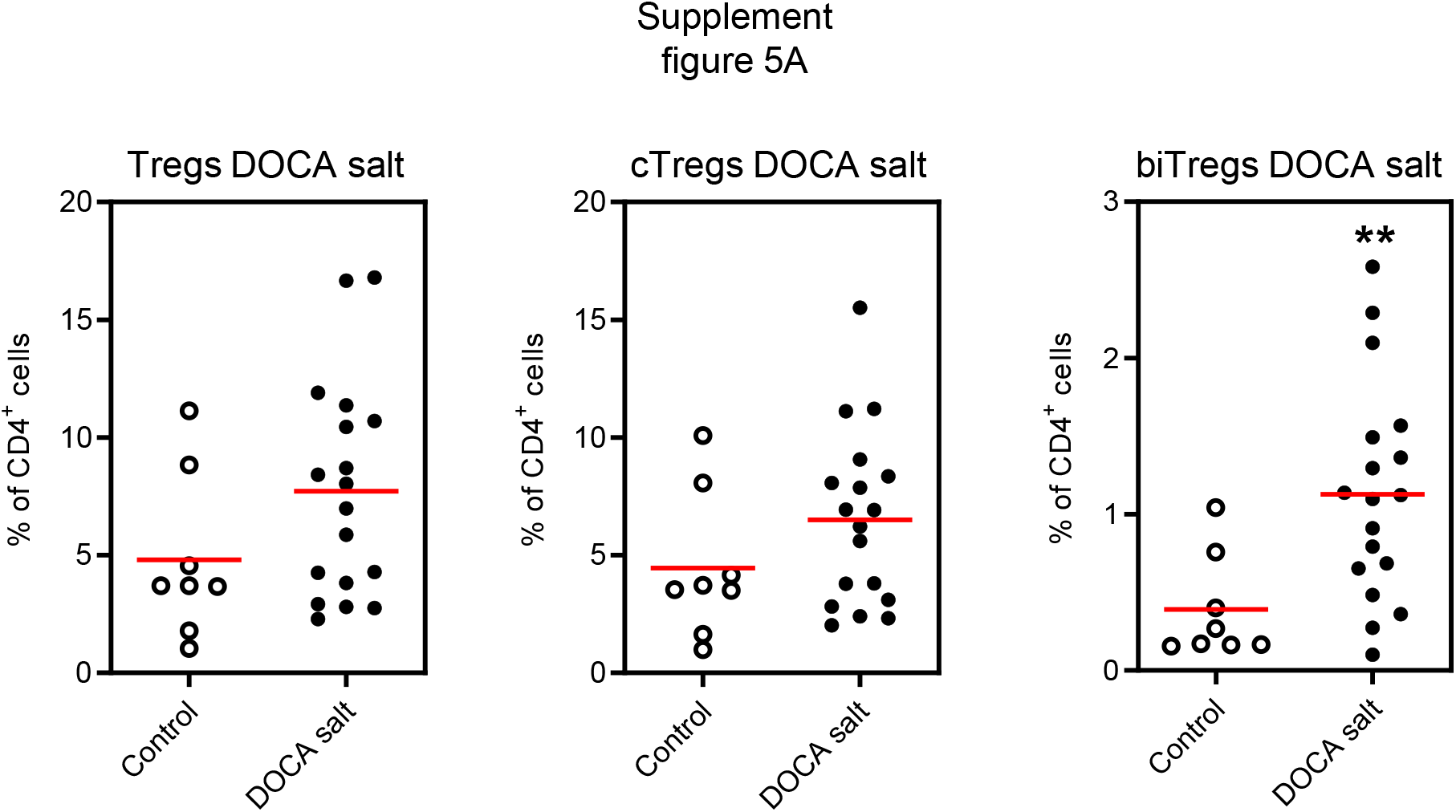
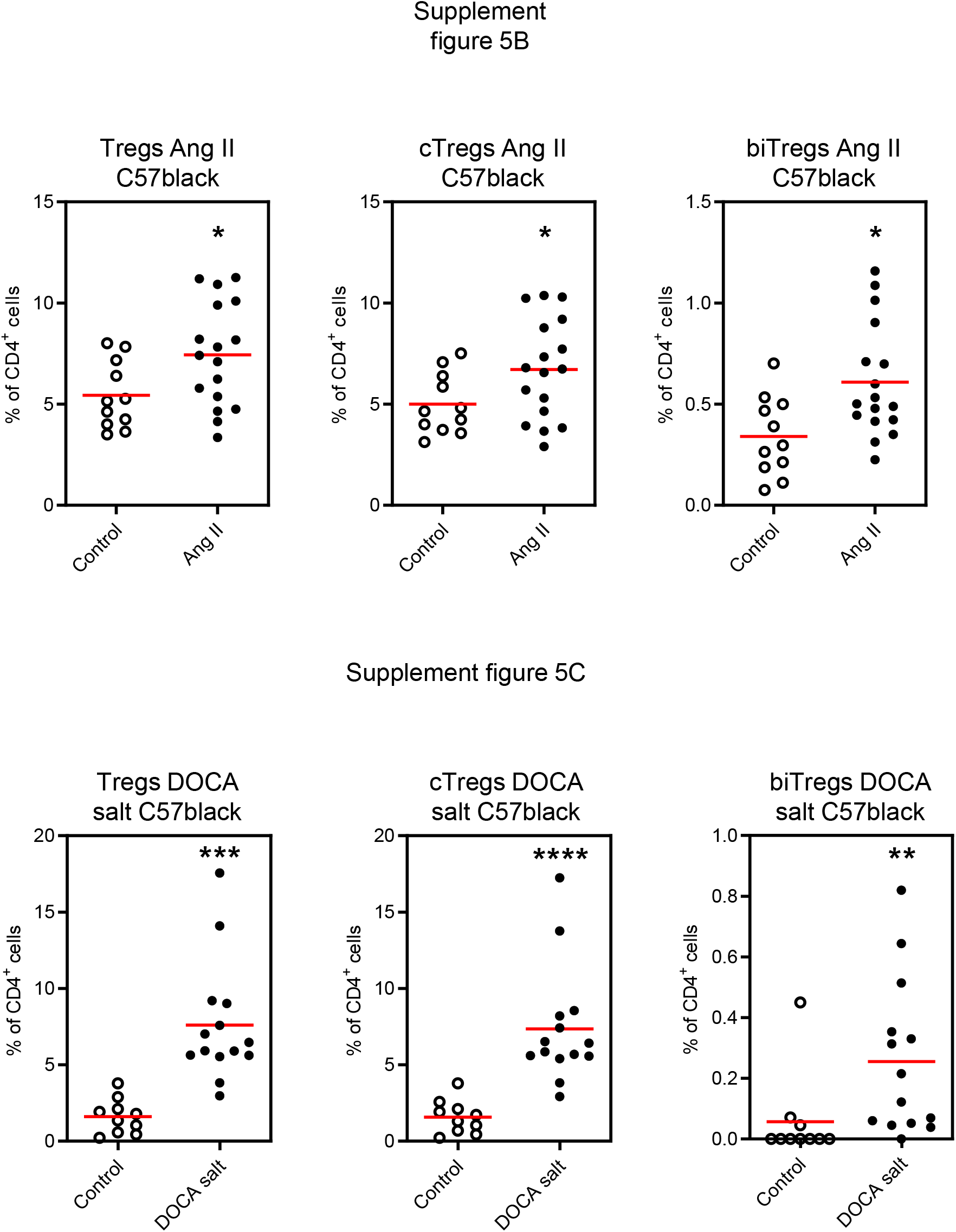
Tregs in experimental hypertension. A: FACS analysis of Tregs infiltrating the kidney in DOCA salt hypertension in BalbC mice. White dots normotensive mice, black dots hypertensive mice. **=p<0.01 vs. control. B: FACS analysis of Tregs infiltrating the kidney in Ang II hypertension in C57black mice. C: FACS analysis of Tregs infiltrating the kidney in DOCA salt hypertension in C57black mice. White dots normotensive mice, black dots hypertensive mice. *=p<0.05, **=p<0.05 ***=p<0.001, ****=p<0.0001 vs. Control.

